# Differing Determinants of Overweight-Obesity and Glucose Intolerance in Offspring Born to Mothers with Diabetes During Pregnancy: Evidence from India

**DOI:** 10.64898/2026.03.25.26349260

**Authors:** Sonali S. Wagle-Patki, Sayali Deshpande-Joshi, Souvik Bandyopadhyay, Sanat Phatak, Shubha Ambardekar, Dattatray Bhat, Deepa Raut, Madhura K. Deshmukh, Rajashree Kamat, Sayali Wadke, Shivani Rangnekar, Rasika Ladkat, Kalyanaraman Kumaran, Pallavi C. Yajnik, Chittaranjan S. Yajnik

**Affiliations:** Kamalnayan Bajaj Diabetology Research Centre, Diabetes Unit, King Edward Memorial Hospital Research Centre, Rasta Peth, Pune, 411011 India; Cytel, Inc, Bengaluru, India; Dr. D.Y. Patil Medical College, Hospital and Research Centre, Dr. D.Y. Patil Vidyapeeth, Pune, India; MRC Lifecourse Epidemiology Unit, University of Southampton.

**Keywords:** bi-parental obesity, obesity and glucose intolerance in offspring of diabetic mothers, maternal hyperglycemia, paternal hyperglycemia

## Abstract

**Background:** Parental diabetes and obesity influence offspring phenotype, but their relative contributions remain unclear.

**Aim:** To examine the relative contributions of parental diabetes and obesity to offspring overweight-obesity and glucose intolerance.

**Methods:** We studied 200 offspring of mothers with diabetes in pregnancy (ODM; 176 indexes, 24 siblings), 176 mothers (133 gestational diabetes (GDM), 22 type 1, 21 type 2 diabetes), and 150 fathers. Controls included 177 offspring of non-diabetic mothers (ONDM), 177 mothers without diabetes in pregnancy, and 163 fathers. Overweight-obesity was defined by WHO criteria, central obesity as waist-to-height ratio >0.5, and glucose intolerance by ADA criteria (fasting glucose for <10 years; oral glucose tolerance test (OGTT) for a≥10 years). Generalized linear mixed-effects models assessed parental determinants of offspring outcomes.

**Results:** ODM were more overweight-obese, centrally obese, and glucose intolerant than ONDM. Younger ODM had higher capillary glucose (5.6 vs 5.1 mmol/L, p<0.001). Among ODM ≥10 years, 37% had prediabetes and 5% diabetes versus 20% and 0% in ONDM. Overweight-obesity was associated with maternal (OR 7.81; 95% CI 2.19-27.85), paternal (OR 6.21; 95% CI 1.57-24.53), and biparental obesity (OR 9.59; 95% CI 2.73-33.69), but not parental diabetes. Glucose intolerance was associated only with maternal diabetes in pregnancy (OR 3.90; 95% CI 2.05-7.41).

**Conclusions:** Preventing offspring obesity will require addressing parental obesity, whereas preventing glucose intolerance will require optimal glycemic control in the mothers before and during pregnancy.

## 1. INTRODUCTION

Maternal diabetes adversely affects pregnancy outcomes and contributes to the epidemic of obesity and diabetes in the young.^1–4^ Both genetic and non-genetic mechanisms, such as epigenetic intrauterine programming and family environment contribute to these outcomes.^5–7^ Classic Native American (Pima Indian) studies showed that maternal hyperglycemia in pregnancy influenced offspring obesity, suggesting a complex interaction of maternal genes and intra-uterine environment, particularly maternal hyperglycemia in influencing offspring’s metabolic health. ^8,9^ In the same population, paternal diabetes also influenced offspring obesity, although the effect was smaller than that of maternal diabetes.^10^ In the Hyperglycemia and Adverse Pregnancy Outcomes (HAPO) studies, maternal gestational diabetes was associated with higher offspring glycemia, lower insulin sensitivity, reduced insulin secretion and disposition index, and a greater risk of obesity at 11-14 years of age, independent of maternal pregnancy BMI. Maternal overweight-obesity is more strongly associated with the child’s obesity-adiposity than GDM (OR 3.0, 2.4-3.7 and OR 1.4, 1.1-1.7, respectively). The HAPO studies also showed a mediating role for birth weight in the association between maternal obesity/glycemia and the child’s adiposity. However, the HAPO did not have any metabolic data from fathers except the history of diabetes.^11–13^ Given the strong link between obesity and diabetes in these populations, it is challenging to separate the effects of these two factors. While the association of maternal diabetes with offspring obesity and glucose intolerance is well established, randomized control trials (RCTs) of intensive control of maternal hyperglycemia during pregnancy had little or no effect on offspring’s long-term obesity-adiposity or glucose intolerance.^14,15^ It is worth noting that these studies were conducted in Western countries, where the pathophysiology of diabetes and obesity differs from that in India.^16^

GDM is common in India, affecting up to 15% of pregnancies.^17^ There are only a few studies that have examined maternal GDM or obesity as a risk factor for offspring outcomes in later in life. A small study of 45 offsprings born to Indian mothers with GDM, showed that adiposity and glucose intolerance were more prevalent in ODMs than in ONDMs during childhood.^18,19^ Given their relatively low BMI compared to their western counterparts, studies in Indian mothers provide a unique opportunity to examine offspring outcomes with discordant maternal exposures. We studied expectant mothers for the last three decades at the Diabetes Unit, King Edward Memorial (KEM) Hospital, Pune, a large tertiary care center, where we found that maternal hyperglycemia rather than BMI was the major driver of offspring adiposity at birth.^20^ For the present case control study, we studied offsprings born to expectant mothers with and without diabetes during pregnancy. We now ask the question: do maternal pregnancy diabetes and overweight-obesity have a distinct influence on offspring’s obesity and glucose intolerance later in life? How does paternal phenotype influence offspring phenotype?

## 2. METHODS AND MATERIALS

### 2.1 Participants

In the present case control study (**In**tergenerational programming of **Dia**besity in offspring of women with **G**estational **D**iabetes **M**ellitus, InDiaGDM)), we reviewed records of 861 pregnancies with diabetes (type 1 diabetes, type 2 diabetes, and GDM) treated in our department over the past 26 years (1988-2014). We contacted these mothers to request the participation of their offspring in this study. Mother, father and their offspring from participating families were requested to participate. They were included in the study after obtaining consent. The anthropometric and biochemical measurements were done between 2014-2017.

### 2.2 Exposures

The diagnosis of diabetes in pregnancy was obtained from medical records. Paternal diabetes was diagnosed by medical history or by a 75-gram OGTT at follow-up based on ADA 2014 criteria. ^21^ Parental overweight-obesity was defined as BMI ≥ 25 kg/m^2^. Parental central obesity was defined as a waist circumference ≥ 85 cm for women and ≥ 90 cm for men. Offspring birth weight was obtained from medical records and converted to a SD score based on the INTERGROWTH-21^st^ study.^22^ We used offspring age, sex at birth, parental smoking status, and standard of living index (SLI), a measure of the family’s socio-economic status (SES)^23^ as covariates in the analysis. Pubertal staging was measured for offspring between 8 to 18 years of age, including date of menarche. ^24^

### 2.3 Control group

As a control group, personnel of the same age group and SES as the ODMs, whose mothers did not have any history of diabetes during pregnancy were included. They were matched for age ±1 year, and sex with ODM. Their parents also served as controls.

### 2.4 Outcomes

#### Offspring small for gestational age (SGA) / large for gestational age at birth (LGA)

we defined SGA as birth weight <10^th^ centile and LGA as birth weight >90^th^ centile of the INTERGROWTH-21^st^ standard.

#### Offspring overweight-obesity

We measured weight and height by a standardized protocol ^25^ (Table S1). We calculated BMI as weight (kg) divided by height (m)^2^. For those aged 2-18 years, overweight-obesity was defined as WHO BMI SD score greater than 1SD, and for those aged 18 years or more, as BMI ≥ 25 kg/m^2^. ^26^

#### Offspring central obesity

Waist circumference was measured using a standardized protocol. ^25^ In the absence of pediatric reference values for central obesity, we used waist-to-height ratio (WHtR) >0.5 to define central obesity. ^27, 28^

#### Offspring glucose intolerance

For offspring younger than 10 years, we performed a single capillary fasting or non-fasting blood glucose measurement on a calibrated glucometer. This was due to the parents’ unwillingness for offspring’s venipuncture. Glucose intolerance for fasting measurements was defined by ADA 2014 criteria of: fasting plasma glucose (FPG) 100 to 125 mg/dl for Impaired Fasting Glucose (IFG); and FPG ≥ 126 mg/dl for diabetes mellitus.

In older offsprings of ≥10 years of age, an OGTT (1.75 grams of anhydrous glucose per kg body weight up to a maximum of 75 grams) was performed, and glucose intolerance was defined based on ADA 2014 criteria (IFG, impaired glucose tolerance: 2 hours OGTT glucose 140 to 199 mg/dl and ≥200 mg/dl as diabetes mellitus). In those already diagnosed with diabetes, we collected a fasting and 2h post prandial serum sample.

### 2.5 Laboratory methods

We measured venous plasma glucose using the glucose oxidase-peroxidase method (Hitachi 902, Roche Diagnostics GmbH, Germany). We compared the performance of the glucometer (Alere Inc., Waltham, MA, United States) against the laboratory plasma glucose measurements on 162 samples. The correlation (r= 0.930, p<0.001) was good across a wide range of concentrations, with a bias of 0.19 mmol/L (95% CI -0.54 - 1.15) such that glucometer readings were higher than venous plasma levels (Figure S1a and 1b). This is within the permissible difference of 0.83 mmol/L provided by the ISO Recommendation for assessing the accuracy for glucometer use in clinical settings.^29^ We measured plasma anti-glutamic acid decarboxylase (GAD65) or anti Zinc Transporter 8 (ZnT8) antibodies using enzyme-linked immunosorbent assay (RSR, Cardiff, UK).

### 2.6 Statistical methods

We calculated the sample size before the study, considering offspring BMI as the primary outcome. The estimated difference in BMI between ODMs and ONDMs was based on previously published literature from India.^18,19^ In the age group less than 10 years, the sample size was calculated to detect 0.5 kg/m^2^ difference between the groups at 90% power and 5% significance level. This amounted to 108 participants in cases and controls. For the older than 10 years age group, the sample size was calculated to detect a 1.0 kg/m^2^ difference between cases and controls at 90% power and 5% significance level, amounting to 77 participants in each group.

We report continuous variables as mean (± SD), if normally distributed, and median (25^th^-75^th^percentiles) if skewed; we report categorical variables as n (%). We tested the difference between the medians of the ODM and ONDM groups by Mann-Whitney test, and the difference in SDs between groups by the independent sample t-test.

We used generalized linear mixed effects models to examine the independent associations between parental obesity and diabetes with offspring overweight-obesity and glucose intolerance. Exposures were parental overweight-obesity at the time of follow-up, categorised as: 1) both parents were of normal weight (reference category: BMI <25 kg/m^2^), 2) only mother was overweight-obese, 3) only father was overweight-obese, and 4) both parents were overweight-obese. Exposures also included parental diabetes status, maternal during pregnancy and paternal at follow-up, categorised as: 1) no diabetes in both parents (reference category), 2) only mother with diabetes during pregnancy, 3) only father with diabetes at follow-up and 4) diabetes in both parents. Offspring birth weight SD score was also included as an exposure. We included a random intercept for the family to account for correlation between siblings. The outcomes included: offspring overweight-obesity, central obesity and glucose intolerance as defined above. Given the long duration of the pregnancy data collection, changes in diagnostic criteria, and advances in diabetes management over time, we generated a categorical year variable dichotomised at the median year of the pregnancy study period (2007), which was included as a covariate in all regression models, like our previous analysis. ^20^ In addition, offspring age, sex, SLI, and parental smoking status were included as covariates. We also show predicted curves to visualize the distribution of offspring BMI, waist to height ratio and glucose across the age, derived from the linear mixed effects models. For these analyses we used following exposures: maternal BMI, paternal BMI and FPG all as continuous variables, and maternal diabetes during pregnancy, as a categorical. We included a random intercept for family to account for correlation among the siblings. Outcomes were offspring BMI SDs, WHtR, and fasting or non-fasting glucose. Offspring age, sex, SLI, and year group (births before or after 2007) were included as covariates.

We used SPSS version 21.0 (IBM corporation, Armonk, NY) and R studio (2026.01.1+403) for statistical analysis. We used *‘lme4’* package for generalized linear mixed effects models and the linear mixed effects models.

### 2.7 Ethics

The study protocol was approved by the Institutional Ethics Committee of the KEM Hospital Research Centre (EC 1333/2014) and is registered with ClinicalTrials.gov (NCT03388723). Participants older than 18 years provided written informed consent. For participants younger than18 years, written parental consent was obtained, and those aged 12-18 years also provided a written assent.

## 3. RESULTS

We reviewed records of 861 mothers who were treated in our diabetes clinic for diabetes during pregnancy and were contacted. We were able to connect with 346 women, and 176 agreed to participate. Among these mothers, 22 had the diagnosis of type 1 diabetes, 21 with type 2 diabetes and 133 with GDM (Figure S2). Women who participated in the study had similar age, BMI, glucose concentrations at diagnosis of GDM, and babies’ birth weight compared to the women who did not participate (all p>0.05, Table S2). We studied 200 ODMs (25 OT1D, 22 OT2D, 153 OGDM), 176 born in the index pregnancy and 24 siblings born in subsequent diabetic pregnancies. We have a wide age range in offspring, of the 200 ODMs studied, 119 were younger than 10 years and 81 were older than 10 years. We also studied 176 mothers and 150 fathers of ODMs. We included 177 ONDMs (93 younger than 10 years and 84 older than 10 years), their mothers (n=177) and fathers (n=163) as controls (Figure S2).

### 3.1 Mothers

Mothers with diabetes in pregnancy in comparison with no diabetes in pregnancy were older (37.4 years vs. 35.1 years), more overweight-obese (67.8% vs 57.4 %), and more centrally obese (52.4% vs 47.6%) at follow-up (Table 1). Among women who had GDM, 24% were classified as prediabetic and 46% as diabetic at follow-up, compared to 27% and 4% respectively in the control group (Table 1). During pregnancy, 25% of the mothers with diabetes were managed by lifestyle modification, 6% were treated with only oral hypoglycemic agents (OHA), 58% were managed with only insulin, and 11% were given both, OHA and insulin.

**Table 1:**
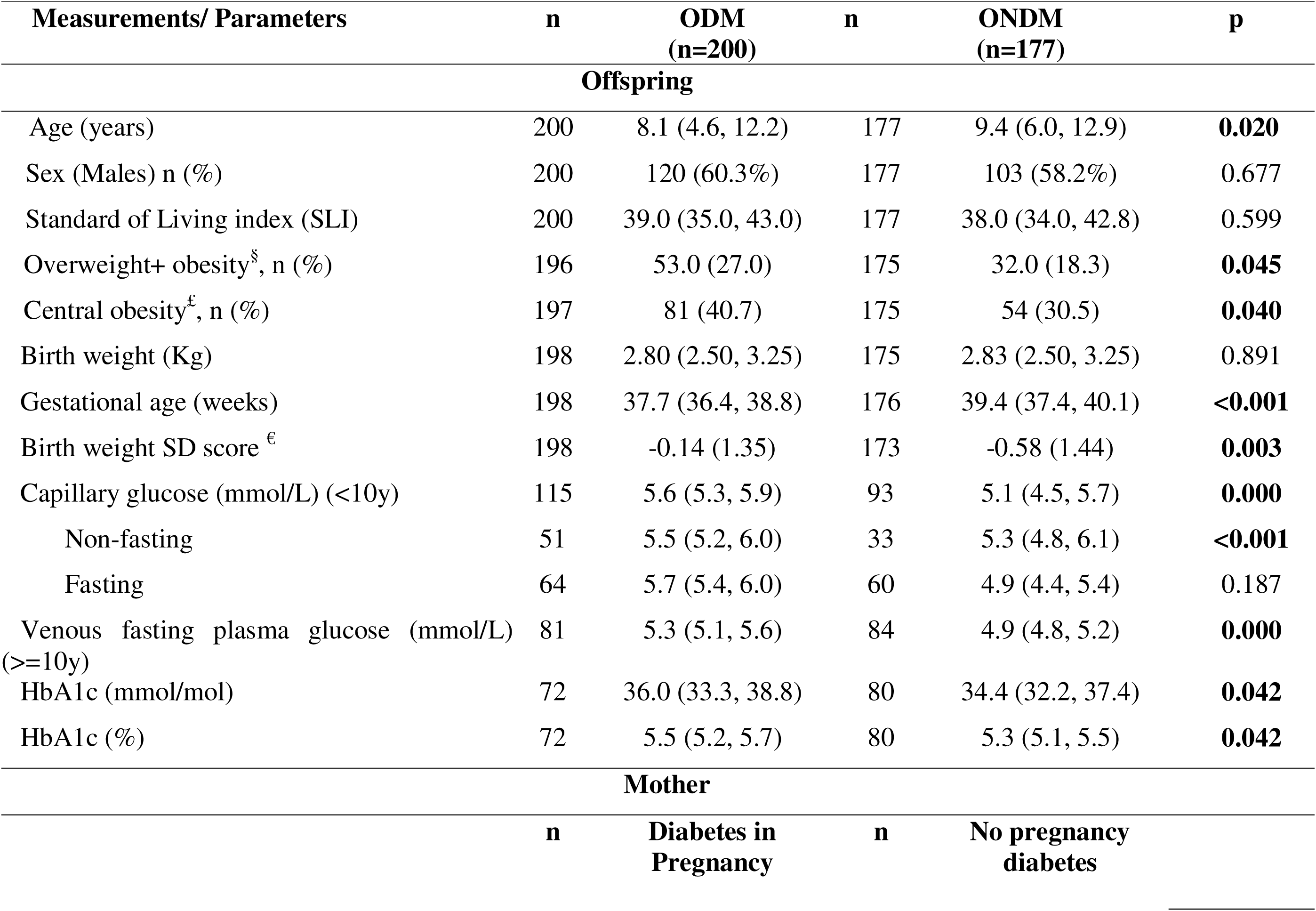
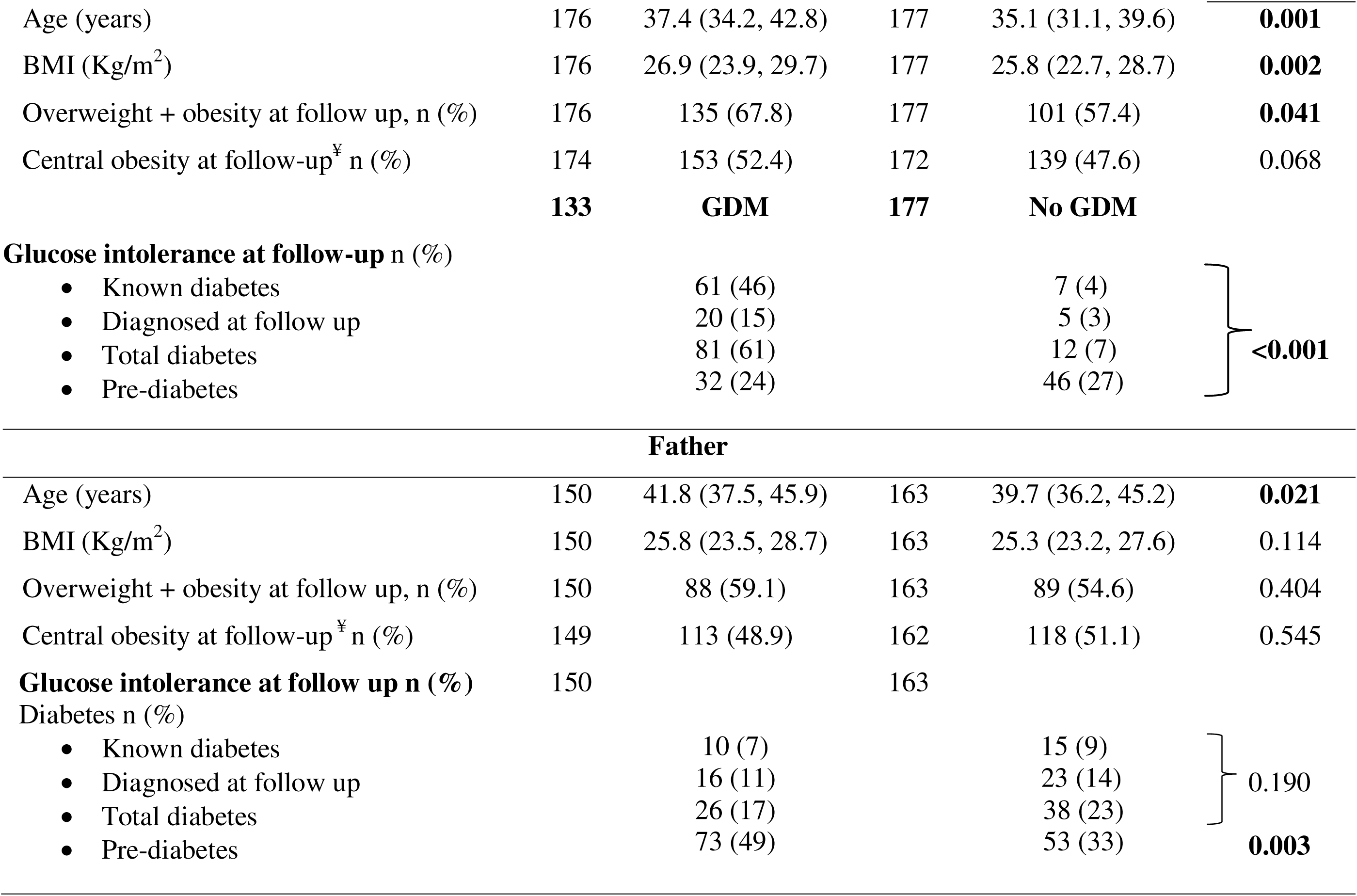
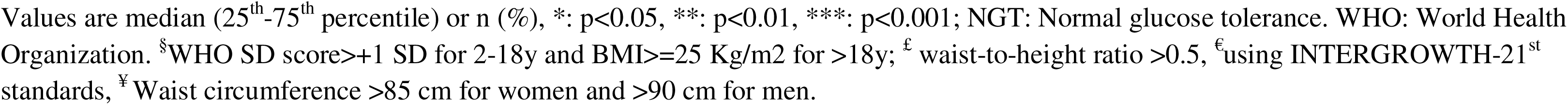
Comparison of body size, composition, and glucose among ODM and ONDM and their parents.

### 3.2 Fathers

Fathers of ODM and ONDM had a similar prevalence of overweight-obesity and central obesity (Table 1). Fathers of ODM had a higher prevalence of pre-diabetes (49% vs 33%, p= 0.03) but similar prevalence of diabetes (17% vs 23%, p=0.190) compared to fathers of ONDM. Hyperglycemic (pre-diabetes + diabetes) fathers were older (median 43.1 years vs 38.4 years, p<0.001) and had a higher BMI (median 26.3 kg/m^2^ vs 24.6 kg/m,^2^ p< 0.001) than normoglycemic fathers.

### 3.3 Offspring

#### 3.3.1 At birth

ODMs were born at an earlier gestation than ONDMs (Table 1). They had higher birthweight SDs but a similar prevalence of LGA (14.1% vs. 8.7%, p=0.101), and lower prevalence of SGA (22.7% vs. 33.7, p=0.021).

#### 3.3.2 At follow-up

ODM and ONDM had similar age, sex, pubertal stage, and SLI (Table 1). Twenty ODM and ONDM females had achieved menarche (at the age of 12.2 ± 1.1 years and 12.2 ± 1.2 years, respectively, p=0.947).

#### 3.3.3 Overweight-obesity and central obesity

ODM had a higher prevalence of overweight + obesity (27.0% vs. 18.3%, p=0.045) and central obesity (40.7% vs. 30.5%, p =0.040) compared to ONDM (Table 1) (Figure 1 A, B) and the difference was more prominent for males (table S4). As a group, the distribution of WHtR was shifted to the right in ODM compared to ONDM (figure S3 B).

**Figure 1:**
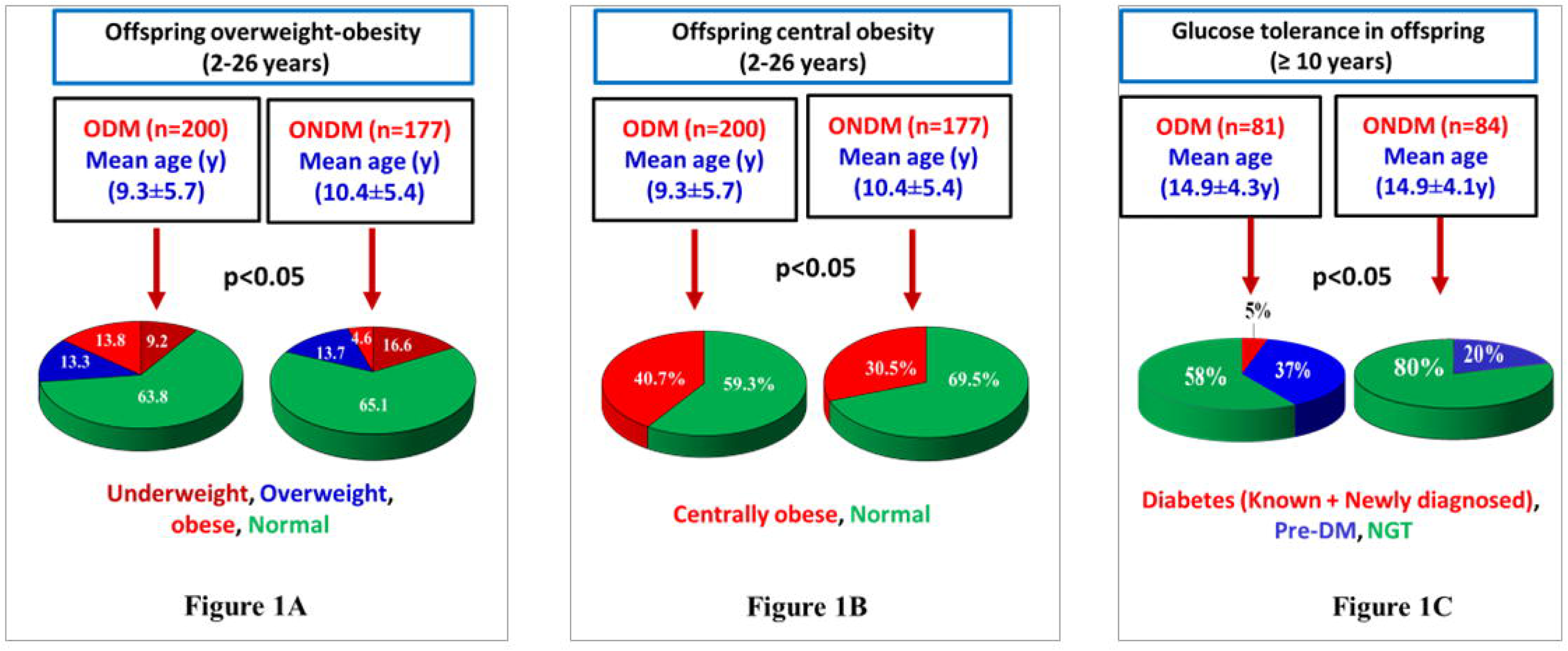
Overweight-obesity, central obesity and glycemic status in offspring. Figure 1A shows prevalence of overweight-obesity in the offspring (2-26 years) using WHO-MGRS, 2007 criteria Figure 1B shows prevalence of central obesity in the offspring (2-26 years) using waist to height ratio > 0.5 Figure 1C shows prevalence of glucose tolerance in the offspring (≥10 years) using ADA 2014 criteria

#### 3.3.4 Glycemia

ODMs had higher glucose and Haemoglobin A1c (HbA1c) concentrations, compared to ONDMs (Table 1, figure S3 C) and the difference was prominent for males (table S4). ODMs <10 years age had higher capillary blood glucose concentrations compared to ONDMs (Table 1). In this group, among those with fasting glucose measurements (n=64), 60.3% ODMs and 21.3% ONDMs (p<0.001) had IFG. Non-fasting capillary glucose measurements were also higher in ODMs compared to ONDMs (Table 1). ODMs ≥ 10 years age had higher prevalence of glucose intolerance on OGTT (5% DM, 37% prediabetes [17.5% IFG, 15% IGT and 5% both IFG and IGT]) compared to ONDM (0% DM, 20% prediabetes [8.3% IFG, 10.7% IGT and 1% both IFG and IGT]) (Figure 1 C). Three ODMs were receiving treatment for diabetes (diagnosed at 14, 16 and 23 years of age); all received OHA, one also received insulin. Based on OGTT, we diagnosed one additional participant with diabetes. All four ODMs with diabetes were clinically classified as type 2 diabetes and were born to GDM mothers. Glucose intolerant ODMs were overweight + obese compared to normal glucose tolerant ODMs (Table S5).

### 3.4 Association of parental overweight-obesity and diabetes with offspring outcomes

#### 3.4.1 Offspring overweight-obesity and central obesity

Using generalized linear mixed effects models, maternal (OR - 7.8, 95% CI 2.2 - 27.8) and paternal (OR - 6.2, 95% CI 1.6 - 24.5) overweight-obesity were directly associated with offspring overweight-obesity and central obesity, when adjusted for offspring’s age, sex, SLI, parental smoking status, and sibling correlation. While offspring’s birthweight was also directly associated with overweight-obesity, parental glycemia was not associated with offspring overweight-obesity or central obesity (Table 2). The additive effect of biparental overweight-obesity on offspring overweight-obesity is seen clearly in Figure S4.

**Table 2:** Generalized linear mixed effects model for factors associated with overweight-obesity, glucose intolerance, central obesity in offspring (ODM + ONDM)

| Exposures | Overweight-obesity<br>(WHO)<br>(n=340) | Central obesity<br>(WHtR >0.5)<br>(n=341) | Glucose intolerance<br>(ADA criteria)<br>(n=264) |
| --- | --- | --- | --- |
| Both parents normal weight at follow up | 1.00 | 1.00 | 1.00 |
| Only mother overweight-obese at follow up | **7.81 (2.19 - 27.85) \*\*** | **3.84 (1.68 - 8.76) \*\*** | 1.84 (0.81 - 4.22) |
| Only father overweight-obese at follow up | **6.21 (1.57 - 24.53) \*\*** | **7.62 (3.14 - 18.51) \*\*\*** | 1.07 (0.41 - 2.82) |
| Both parents overweight-obese at follow up | **9.59 (2.73 - 33.69) \*\*\*** | **4.85 (2.16 - 10.90) \*\*\*** | 1.09 (0.49 - 2.42) |
| No parent with diabetes | 1.00 | 1.00 | 1.00 |
| Only mother with diabetes in pregnancy | 1.57 (0.81 - 3.04) | 1.47 (0.86 - 2.51) | **3.90 (2.05 - 7.41) \*\*\*** |
| Only father with diabetes at follow up | 1.53 (0.60 - 3.89) | 1.04 (0.46 - 2.37) | 1.04 (0.39 - 2.78) |
| Both parents with diabetes | 1.48 (0.53 - 4.14) | 1.34 (0.56 - 3.20) | **3.02 (1.11 - 8.20) \*** |
| Offspring birthweight SD score | **1.27 (1.03 - 1.57) \*** | 1.13 (0.95 - 1.34) | 1.20 (0.98 - 1.46) |

Results expressed as Odds ratio (95% Confidence intervals). Significant results are shown in bold. Exposures included: Parental overweight-obesity ( $\text{BMI} \geq 25 \text{ Kg/m}^2$ ) at follow up, glucose intolerance in father at follow up (known diabetes + newly diagnosed, using ADA 2014 OGTT criteria) and maternal diabetes during pregnancy (yes or no), offspring birth weight (SD score of birth weight calculated using INTERGROWTH-21<sup>st</sup> standards). Outcomes included: 1. Overweight-obesity in offspring based on BMI (WHO), 2. Central obesity in offspring defined as : waist-to-height ratio $>0.5$ . 3. Glucose intolerance in offspring: for those younger than 10 years- IFG based on fasting capillary glucose using ADA 2014 criteria; for those older than 10 years - known diabetes + newly diagnosed based on OGTT glucose using ADA 2014 criteria. We included a random intercept for family to account for the correlation among the siblings. Covariates included a categorical year variable dichotomised at the median year of the study period (2007), offspring's age, sex, SLI, parental smoking status.

#### 3.4.2 Offspring glucose intolerance

Maternal diabetes during pregnancy predicted offspring’s glucose intolerance (OR- 3.9, 95% CI 2.0 - 7.4), adjusting for offspring’s age, sex, SLI, parental smoking status and sibling correlation in generalized linear mixed effects models. Father’s diabetes or parent’s overweight-obesity were not predictive (Table 2).

Unlike for overweight-obesity, an additive effect of biparental diabetes on offspring glucose intolerance was not observed.

### 3.5 Predictive modelling of offspring BMI, WHtR, and glucose measurements

Predicted BMI and WHtR from the linear mixed effects models were plotted against age (Figure 2 A, B). BMI curves are reminiscent of the classic description of a catch-down in infancy, an earlier adiposity-rebound in childhood and more rapid growth subsequently in ODM compared to the ONDMs. Predicted curves for glucose showed consistently higher glucose concentrations from early childhood through adulthood in ODMs compared to ONDMs (Figure 2 C, D).

**Figure 2:**
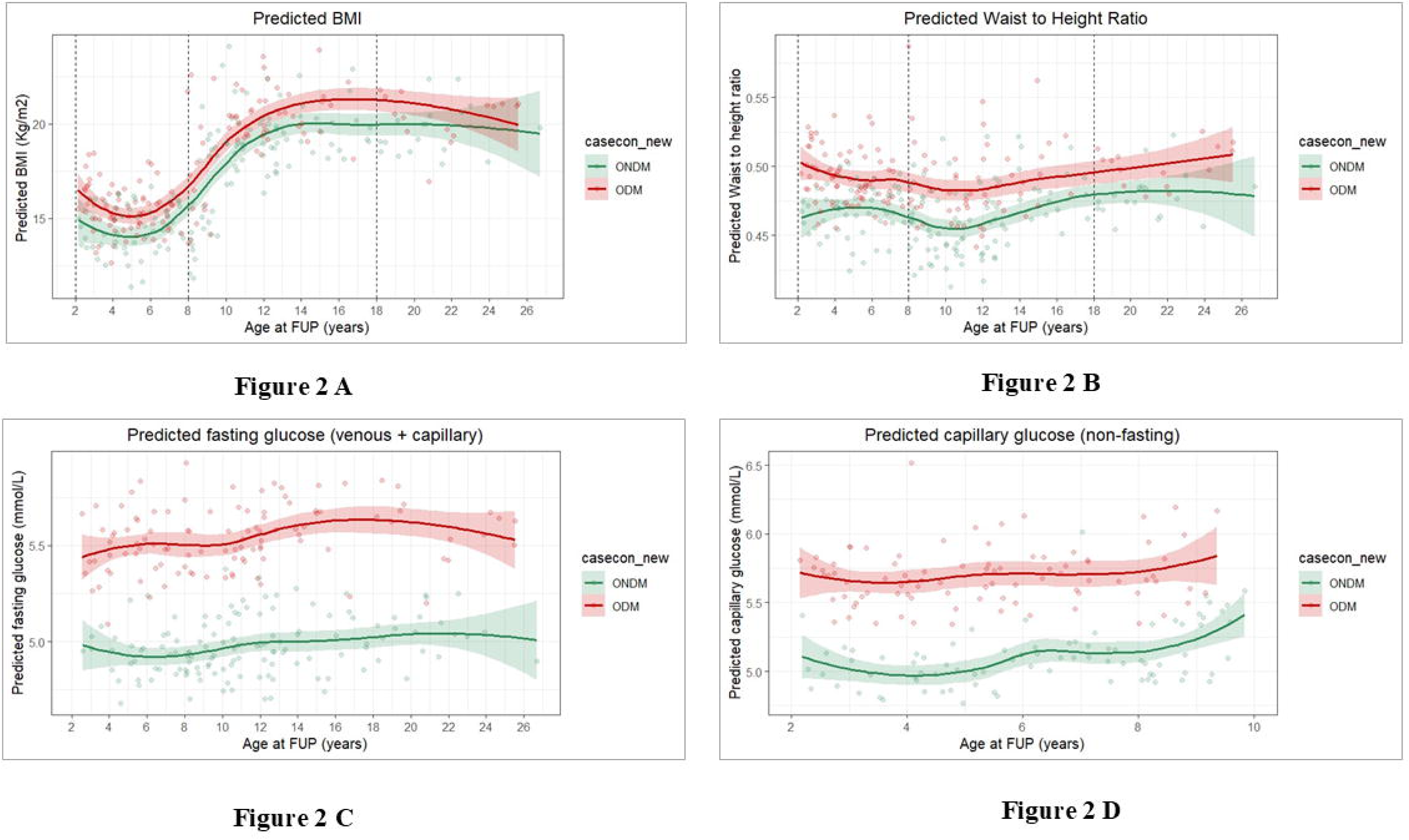
Predicted body mass index (BMI), waist circumference, and glucose concentrations in ODM (shown in red) and ONDM (shown in green) Figure 2 A shows predicted curves for BMI, figure 2B shows predicted curves for waist-to-height ratio, 2C shows predicted curves for fasting glucose concentrations (both, capillary and venous), 2D shows predicted curves for capillary glucose (non-fasting) in offspring younger than 10 years

### 3.6 Maternal diabetes type and offspring phenotypes

Mothers with type 1 diabetes had lowest BMI, those with GDM intermediate and those with type 2 diabetes were the most overweight-obese (Table S3 and Figure S4). Maternal BMI reflected in offspring BMI, children of mothers with type 1 diabetes had the lowest BMI and those of mothers with type 2 diabetes had the highest BMI. ODMs born to mothers with all three diabetes types had higher glucose concentrations compared to ONDMs. Type of treatment for maternal diabetes in pregnancy, whether it was lifestyle management, OHA or insulin was not related to the outcome. None of the ODMs had positive antibodies for GAD65 or Zn-T8.

### 3.7 A model for intergenerational transmission of adiposity and glucose intolerance

Based on these results, and data from previous publication,^20^ we propose a model for neonatal and later life outcomes in the ODMs. Figure 3 demonstrates that maternal hyperglycemia results in fetal hyperinsulinemia, which promotes fetal adiposity and macrosomia. Intrauterine exposure to excess insulin can lead to later-life metabolic dysfunction, leading to glucose intolerance in offspring. Bi-parental obesity (additive effect) leads to offspring obesity in later life, partially mediated by larger birth size.

**Figure 3:**
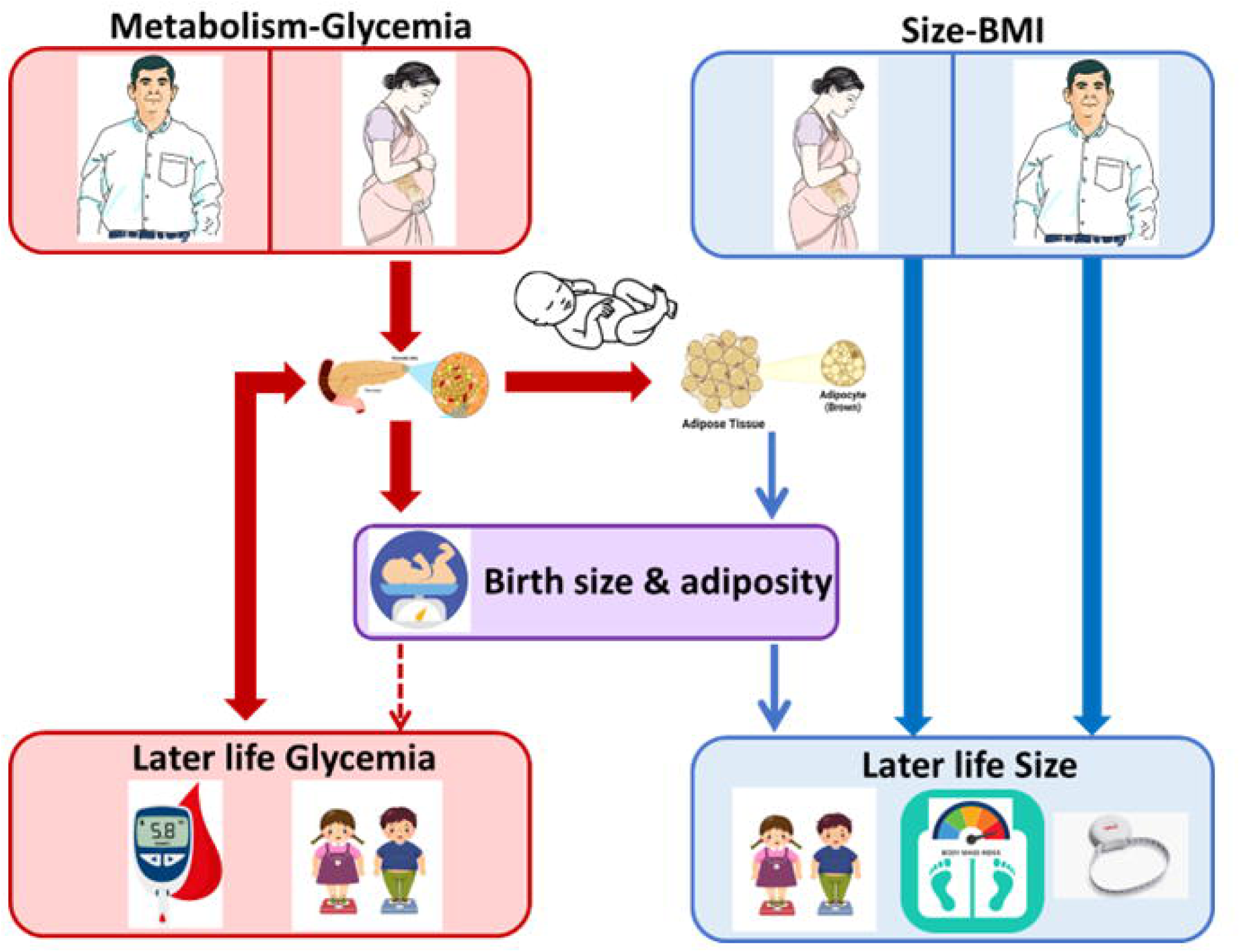
Intergenerational transmission of adiposity and glucose intolerance. Figure shows that maternal hyperglycemia stimulates fetal hyperinsulinism which promotes adiposity and macrosomia. Intra-uterine beta-cell hyperstimulation programs later life dysfunction, contributing to glucose intolerance. Offspring’s later life obesity is influenced predominantly by bi-parental obesity (additive) and to a smaller extent by larger birth size. Genetic, epigenetic, and family environment will influence these complex mechanisms.

## 4. DISCUSSION

Our results in Indian children demonstrate that those born in pregnancies with diabetes (ODM) have higher BMI, WHtR, and glycemia compared to those born in pregnancies without diabetes (ONDM). This reflected in higher prevalence of overweight-obesity, central obesity and glucose intolerance in ODMs. These phenotypes were apparent from early childhood and persisted into adulthood. The predicted BMI curve (Figure 2A) is suggestive of a catch-down in infancy, an earlier adiposity-rebound in childhood, and a subsequently more rapid growth in ODM compared to ONDM. Curves for glucose showed consistently higher glucose concentrations in the ODM as compared to ONDMs across the age range (Figure 2C and 2D). The risk of overweight-obesity and central obesity in offspring was associated with bi-parental overweight-obesity in an additive manner, but not with parental diabetes. On the other hand, glucose intolerance in the offspring was entirely predicted by maternal diabetes in pregnancy, and not by paternal diabetes or parents’ overweight-obesity, highlighting a primary role for maternal glycemic status during pregnancy. In our previous study, we have reported a primary role for maternal diabetes rather than her obesity for neonatal adiposity.^20^ Thus, the two related but distinct parental characteristics (obesity and diabetes) have differing influences on offspring phenotype during different stages in the life course, suggesting a complex interplay of genetic, and epigenetic factors influenced by intrauterine and family environment.

### 4.1 Offspring overweight-obesity

Studies among pregnancies without diabetes show that parental obesity, both maternal and paternal, is a major determinant of offspring obesity.^30–32^ Among pregnancies complicated by diabetes, most studies report a higher prevalence of obesity in offspring compared with those born to women without diabetes.^2–4,8–10,18,19^ In the HAPO study, maternal gestational glycemia and BMI were independently and additively associated with childhood adiposity,^33^ with birth weight acting as a mediator.^34^ Some studies have also reported an additional contribution of paternal obesity.^35–38^ In Western populations, where obesity and diabetes frequently coexist, disentangling their independent effects is challenging.^10^

In our population, offspring overweight-obesity and central obesity were associated with parental overweight-obesity, but not with their diabetes. Thus, offspring of the mothers with lowest BMI (type 1 diabetes) had the lowest BMI, and those of mothers with highest BMI (type 2 diabetes) had the highest BMI. Mothers with type 1 diabetes were the most hyperglycemic, those with type 2 diabetes, less so. This dissociation of maternal obesity and hyperglycemia enabled appreciation of their specific influences on offspring phenotype. The additive effect of paternal obesity suggests genetic and/or shared environmental contributions. Our observations are largely consistent with previous reports,^39, 40^ except for the Kaiser Permanente study,^41, 42^ where maternal obesity was substantially higher than in our cohort.

Longitudinal studies have shown that ODM exhibit catch-down growth in infancy, early adiposity rebound, and excess growth later in childhood compared with ONDM. ^18,19, 43–46^ Our predicted BMI and WHtR trajectories in cross sectional data demonstrated a similar pattern (Figure 2A). Sex-specific effects have also been reported. In the Parthenon study ^18,19^ and the Hong Kong HAPO study,^47^ females were predominantly affected, whereas males were more affected in our cohort. Most Western studies report either similar adiposity across sexes or a male excess.^48^

### 4.2 Offspring glucose intolerance

In our study, ODM had higher glycemia than ONDM from early childhood into young adulthood. Early-onset type 2 diabetes (<20 years) occurred exclusively in ODM, and prediabetes was substantially more prevalent. Offspring glucose intolerance was entirely predicted by maternal diabetes during pregnancy, irrespective of diabetes type, and was unrelated to paternal diabetes or parental overweight-obesity. This highlights a dominant role of maternal hyperglycemia. Notably, offspring of mothers with type 1 diabetes manifested features of type 2 rather than autoimmune diabetes, indicating that intrauterine hyperglycemia, rather than immunogenetics, underlies this risk. Thus, maternal diabetes during pregnancy was the principal determinant of offspring glucose intolerance, in contrast to the pattern observed for offspring overweight-obesity.

The increased risk of diabetes in ODM was first demonstrated in studies from Chicago and among Native Americans (Pima Indians).^2–4,10,49,50^ Pedersen’s^51^ hyperglycemia-hyperinsulinemia hypothesis and Freinkel’s^52^ fuel-mediated teratogenesis concept proposed that intrauterine overnutrition in diabetic pregnancies programs fetal metabolism, predisposing to obesity and diabetes later in life. This paradigm was validated by the HAPO study, which showed a direct association between in utero exposure to maternal hyperglycemia and offspring glucose intolerance.^11–13^ A Danish study reported an eight-fold higher risk of type 2 diabetes/prediabetes in offspring of GDM and a four-fold higher risk in offspring of mothers with type 1 diabetes compared with the background population, independent of birth weight.^50^

Studies among Pima Indians further demonstrated a higher diabetes risk in siblings born after maternal diabetes onset compared with those born before, with no such effect observed for paternal diabetes, supporting a predominant intrauterine influence over genetic transmission.^10^ Consistent with the Developmental Origins of Health and Disease (DOHaD) paradigm, epigenetic programming by the intrauterine environment likely contributes to this risk.^53–56^ The Pune Maternal Nutrition Study demonstrated an intergenerational association between maternal glycemia and offspring glucose intolerance at 18 years, even when maternal glucose levels were within the normal range during pregnancy.^54^ A parent-of-origin effect was also observed, shifting from paternal to maternal influence with increasing offspring age.^55^ Finally, recent HAPO follow-up data showed an additive effect of genetic susceptibility and in utero exposure to maternal hyperglycemia on the risk of glucose intolerance (IGT and type 2 diabetes).^56^

### 4.3 Intergenerational transmission of obesity and glucose intolerance

Based on our findings, we have constructed a mechanistic model to show transmission of overweight-obesity and diabetes risk from the parents to the offspring (Figure 3). We suggest that maternal diabetes targets fetal beta cells, causing hyperinsulinemia and overgrowth of insulin-sensitive tissues (especially adipose tissue).

However, later life overweight-obesity, was driven by bi-parental overweight- obesity in an additive manner. The latter could be due to genetic factors and/or family environment. This simplistic scheme may help public health interventions. Offspring diabetes may be prevented by strict maternal glycemic control before and during pregnancy, while overweight-obesity may be controlled by targeting the family environment.

### 4.4 Strengths and limitations

We report follow-up of the largest cohort of Indian offspring born to mothers with diabetes, spanning a wide age range and encompassing different types of maternal diabetes. The relative thinness (lower BMI) of mothers with diabetes compared to the Western populations and contrasting phenotypes of mothers with type 1, type 2 and GDM provided an opportunity to disentangle the effects of maternal obesity and glucose intolerance on corresponding offspring phenotypes. Inclusion of fathers allowed assessment of paternal contributions to a condition traditionally attributed to maternal diabetes, although paternal measurements were available only at follow-up and not during mother’s pregnancy.

Unlike previous studies with narrow offspring age ranges, our study spans a wide age range among ODM, demonstrating that abnormalities in body size and metabolism emerge in early childhood and persist into later life. Interestingly, our predictive models based on cross-sectional data showed a catch-down growth in infancy and an earlier adiposity rebound in ODM compared with ONDM for BMI, reminiscent of curves based on longitudinal growth studies. In offspring younger than 10 years, only capillary glucose measurements were feasible for ethical and pragmatic reasons. Glucometer values showed good agreement with laboratory measurements, with bias within acceptable analytical limits. Only one-fifth of eligible women could be followed up; however, pregnancy characteristics of mothers and offspring were similar between those who participated in the study and those who did not, supporting representativeness. As a hospital-based study, some selection bias is possible, although KEM Hospital serves individuals across all socioeconomic strata.

Diagnostic criteria and management of diabetes in pregnancy evolved during the study period; this was addressed statistically by inclusion of a categorical year variable and the associations persisted after adjustment. The control group was defined based on maternal recall of GDM, but the low prevalence of glucose intolerance at follow-up suggests minimal misclassification. We lacked data on pregnancy HbA1c, gestational weight gain, and breastfeeding. Although lifestyle information including diet and physical activity was collected, associations with the outcomes will be reported separately. Our findings are generalizable to Indian populations; generalizability to other populations will be known when similar analyses are performed in other studies.

In summary, we confirm an increased long-term risk of overweight-obesity, central obesity, and glucose intolerance in Indian ODMs. We show that overweight-obesity and central obesity are driven by bi-parental overweight-obesity, and glucose intolerance by maternal diabetes in pregnancy. We have previously shown that neonatal adiposity is influenced primarily by maternal diabetes, and our model thus highlights the contrasting origins of intrauterine and later life adiposity. Strict maternal glycaemic control before and during pregnancy will help limit fetal overgrowth and subsequent glucose intolerance, while family-based lifestyle interventions will help reduce childhood obesity and adiposity. Genetic, epigenetic, and environmental determinants of these observations warrant further investigation and could provide insights into molecular mechanisms and targets for intervention.

## Supporting information

Online supporting information - ODM

## AUTHOR CONTRIBUTIONS

SWP, KK, MD, SA, and CSY were involved in study conceptualization, participant recruitment, and actual conduct of the study. SWP, RL, PCY were involved in study administration. SWP, SDJ and SB were involved in data curation and formal analysis. DB, DR, RK, SW and SR were involved in laboratory analysis. SWP, SDJ, SP and CSY were involved in data visualization, writing, and reviewing the manuscript. All authors critically reviewed and approved the final version.

## ACKNOWLEDGEMENTS

We are grateful to Prof. Edwin Gale, Prof. Caroline Fall, and Dr. Arijeet Gattu for discussions. We also acknowledge Dr. Shailaja Kale, Dr. Smita Dhadge, Dr. Meenakumari, Neelam Memane, Aboli Bhalerao, Swati Alekar, Alma Baptist, Preeti Kalel, Ajay, Abhay, Dr. Rohan Shah, Rupali Joshi for their valuable contributions to the study.

## DATA ACCESSIBILITY STATEMENT

Data is available from Prof. C. S. Yajnik by applying with a 200-word plan of analysis. Data sharing is subject to KEM Hospital Research Centre Ethics Committee approval and Govt. of India’s Health Ministry Advisory Committee permission.

## DECLARATION OF GENERATIVE AI AND AI-ASSISTED TECHNOLOGIES IN THE WRITING PROCESS

We have used ChatGPT to generate human figures (used in the graphical abstract) and to assist with basic language editing (grammar and spelling) in selected paragraphs. The authors reviewed and edited all content after using this tool.

## DECLARATION

Aspects of this work have been presented at the following international DOHaD 2013, Singapore; 12th International Congress on Obesity 2014, Malaysia; DOHaD 2017, The Netherlands; ADA 2018, USA; ADA 2019, USA; DOHaD 2019, Australia; DOHaD 2022, Canada; ICOPH 2024 - 10th International Conference on Public Health 2024,Thailand; and national conferences: SNEHA 2014, DIPSI 2016, SNEHA 2017, NSI 2018, RSSDI 2021, ACCS 2023 (ACCS-9), SNEHA 2023, RSSDI 2023, Regional Young Investigators’ Meeting, 2024, NSI 2024 India.

## Abbreviations

ODM: Offspring of mother with diabetes
GDM: Gestational Diabetes Mellitus
ONDM: Offspring of mother without diabetes
BMI: Body Mass Index
WHO: World Health Organization
SDs: Standard Deviation scores
OGTT: Oral Glucose Tolerance Test
OR: Odds Ratio
CI: Confidence Interval
HAPO: Hyperglycemia and Adverse Pregnancy Outcomes
RCTs: Randomized Control Trials
KEM Hospital: King Edward Memorial Hospital
InDiaGDM: **In**tergenerational programming of **Dia**besity in offspring of women with **G**estational **D**iabetes **M**ellitus
ADA: American Diabetes Association
SLI: Standard of Living Index (SLI)
SES: Socioeconomic status
SGA: Small for Gestational Age
LGA: Large for Gestational Age
WHtR: Waist-to-Height Ratio
FPG: Fasting Plasma Glucose
IFG: Impaired Fasting Glucose
IGT: Impaired Glucose Test
GAD: anti Glutamic Acid Decarboxylase
ZnT8: anti Zinc Transporter 8 (ZnT8)
OHA: Oral Hypoglycemic Agents
HbA1c: Haemoglobin A1c
DOHaD: Developmental Origins of Health and Disease
NGT: Normal Glucose Tolerant

