## Supplementary material for "Differing Determinants of Overweight-Obesity and Glucose Intolerance in Offspring Born to Mothers with Diabetes During Pregnancy: Evidence from India": Online supporting information - ODM

**Address:**

Diabetes Unit,

6th floor, Banoo Coyaji Building,

KEM Hospital Research Centre,

Maharashtra, India

**Contact number:** +919822847481

**Table S1: Methodology of anthropometric measurements**

|  | Site of measurement | Instrument and make |  |  | | |
| --- | --- | --- | --- | --- | --- | --- |
|  |  |  | Least count | Offspring | Mother | Father |
| Weight | - | Electronic weighing scales  (ATCO Healthcare Ltd, Mumbai, India) | 0.01 kg | ✓ | ✓ | ✓ |
| Height | Head (Frankfurt plane) | Stadiometer (CMS Instruments Ltd, London, UK). | 0.1 cm | ✓ | ✓ | ✓ |
| -BMI | Weight (Kg)/ (height, m^2^) | - | - | ✓ | ✓ | ✓ |
| Waist circumference | Mid-point between lower border of the costal margin and the upper border of the iliac crest in the mid-axillary line | Non- stretchable fiberglass tape (CMS Instruments, London, UK) | 0.1 cm | ✓ | ✓ | ✓ |
| Hip circumference | The greater trochanter (the widest portion of the hip) | Non- stretchable fiberglass tape (CMS Instruments, London, UK) | 0.1 cm | ✓ | ✓ | ✓ |
| Skinfold thickness | |  |  |  |  |  |
| Triceps | The posterior most bulging portion over the tricep at  the mid-point between tip of  shoulder and tip of olecranon process on the non- dominant arm | Harpenden skinfold calipers (CMS Instruments, London, UK) | 0.1 mm | ✓ | X | X |
| Biceps | The anterior most bulging portion of upper arm over biceps at the mid-point between tip of shoulder and tip of olecranon process on the non-dominant arm | Harpenden skinfold calipers (CMS Instruments, London, UK) | 0.1 mm | ✓ | X | X |
| Subscapular | Immediately below the inferior angle of the scapula | Harpenden skinfold calipers (CMS Instruments, London, UK) | 0.1 mm | ✓ | X | X |
| Suprailliac | Upper border of the iliac crest is palpated and marked with a pen in the mid-axillary line on the non-dominant side | Harpenden skinfold calipers (CMS Instruments, London, UK) | 0.1 mm | ✓ | X | X |

**Table S 2: Characteristics of the mothers who were followed and not followed up in later life study (InDiaGDM (Arm-3) study)**

|  | **Followed (n=176)** | **Not followed (n=685)** | p |
| --- | --- | --- | --- |
| **Mothers** | | | |
| Age at conception (years) | 29.4 (4.7) | 29.5 (4.6) | 0.827 |
| Height (cm) | 154.3 (6.2) | 154.9 (6.3) | 0.433 |
| BMI (Kg/m^2^) | 23.5 (21.3-26.6) | 24.6 (22.2-27.3) | 0.134 |
| FPG (mmol/L) | 5.4 (4.8-6.7) | 5.7 (4.8-6.8) | 0.484 |
| 2-hour glucose (mmol/L) | 9.1 (7.8-11.3) | 9.4 (8.2-11.2) | 0.379 |
| **Offspring** | | | |
| Birth weight (gm) | 2821.4 (641.3) | 2897.8 (639.5) | 0.191 |

Values are mean, (SD) for normally distributed variables or median (25^th^ and 75^th^ percentiles) for skewed variables. BMI: Body mass index, FPG: Fasting plasma glucose.

### **Table S 3: Parental and offspring measurements in later life study according to maternal type of diabetes in pregnancy**

|  | **Maternal measurements at follow-up** | | | | | | | | |
| --- | --- | --- | --- | --- | --- | --- | --- | --- | --- |
| **Measurements** | **No diabetes**  **(n=177)** | **Type 1 diabetes (n=22)** | **Type 2 diabetes**  **(n=21)** | **Gestational**  **Diabetes**  **(n=133)** | **All diabetes**  **(n=176)** | **p1** | **p2** | **p3** | **p4** |
| Age (years) | 35.1 (31.1-39.6) | 32.8 (30.5-40.9) | 37.3 (34.5-38.9) | 38.4 (34.5-44.2) | 37.4 (34.2-42.8) | 1.000 | 1.000 | **<0.001** | **0.001** |
| Height (cm) | 155.0 (151.5-158.2) | 157.5 (150.9-162.4) | 154.8 (150.4-159.2) | 154.2 (149.8-156.2) | 154.5 (150.5-157.5) | 1.000 | 1.000 | 0.245 | 0.210 |
| Weight (kg) | 60.7 (54.1-70.2) | 55.1 (52.7-61.9) | 68.9 (62.7-84.4) | 62.8 (56.7-71.6) | 62.8 (56.7-71.5) | 1.000 | **0.017** | 0.066 | **0.018** |
| BMI (Kg/m^2^) | 25.8 (22.7-28.7) | 22.9 (22.1-25.3) | 28.7 (27.2-32.7) | 27.1 (23.9-30.6) | 26.9 (23.9-29.7) | 0.427 | **0.013** | **0.002** | **0.002** |
| Waist (cm) | 89.1 (82.3-97.3) | 83.7 (81.3-91.0) | 100.8 (94.5-107.1) | 91.0 (85.7-98.7) | 91.2 (85.0-98.7) | 0.837 | **0.001** | **0.003** | **0.001** |
| Hip (cm) | 101.4 (95.0-107.9) | 98.0 (97.0-100.5) | 107.5 (101.3-118.0) | 103.0 (95.2-111.2) | 102.4 (96.3-110.8) | 1.000 | **0.001** | **0.003** | **<0.001** |
| Waist hip ratio | 0.88 (0.83-0.91) | 0.85 (0.83-0.89) | 0.92 (0.89-0.97) | 0.89 (0.86-0.93) | 0.89 (0.85-0.94) | 1.000 | **<0.001** | **0.003** | **<0.001** |
| Central obesity, n (%) (Waist hip ratio >0.8) | 149 (84.1) | 18 (81.8) | 17 (80.9) | 126 (94.7) | 161 (92.0) | 0.776 | 0.703 | **0.003** | 0.105 |
| Overweight+obesity, n (%) | 101 (57.4) | 8 (36.4) | 17 (81.0) | 91 (68.9) | 135 (67.8) | 0.062 | **0.037** | **0.038** | **0.041** |
|  | **Offspring measurements** | | | | | | | | |
| **Measurements** | **No diabetes**  **(n=177)** | **Type 1 diabetes (n=25)** | **Type 2 diabetes**  **(n=22)** | **Gestational**  **Diabetes**  **(n=153)** | **All diabetes**  **(n=200)** | **p1** | **p2** | **p3** | **p4** |
| Age (years) | 9.4 (6.0-12.9) | 6.9 (4.4-15.3) | 5.1 (3.7-7.6) | 8.8 (5.1-12.9) | 8.1 (4.6-12.2) | 0.100 | **0.048** | 0.451 | 0.963 |
| SLI | 38.0 (34.0-42.8) | 37.5 (31.3-42.3) | 38.0 (34.0-41.0) | 40.0 (35.0-43.5) | 39.0 (35.0-43.0) | 0.449 | 0.399 | 0.223 | 0.464 |
| Birthweight (Kg) | 2.83 (2.50-3.25) | 2.70 (2.37-3.16) | 2.51 (2.46-3.09) | 2.85 (2.52-3.30) | 2.80 (2.50-3.25) | 0.124 | 0.050 | **0.018** | **0.047** |
| Birthweight SD score | -0.15 (-0.82-0.69) | 0.16 (-0.60-0.55) | -0.23 (-0.84-0.53) | 0.11 (-0.48-0.89) | 0.11 (-0.53-0.79) | 0.158 | 0.066 | **0.027** | 0.066 |
| Height (cm) | 132.8 (113.6-152.9) | 114.9 (104.7-152.7) | 109.5 (97.7-124.2) | 131.6 (106-8-154.6) | 126.8 (105.1-152.9) | 0.237 | 0.305 | 0.181 | 0.489 |
| Weight (Kg) | 28.4 (17.9-45.2) | 20.0 (16.2-43.8) | 15.8 (12.8-27.9) | 29.8 (17.1-48.8) | 26.5 (15.8-45.9) | 0.058 | 0.068 | **0.022** | 0.095 |
| BMI (Kg/m^2^) | 16.2 (14.1-19.1) | 15.5 (13.9-18.1) | 14.3 (13.4-18.2) | 16.8 (14.5-21.1) | 16.2 (14.4-20.8) | **0.001** | **0.000** | **0.000** | **0.000** |
| Overweight+obesity, n (%) | 25 (14.2) | 3 (12) | 5 (22.7) | 39 (26) | 47 (23.9) | 0.765 | 0.293 | **0.007** | **0.018** |
| Waist (cm) | 59.8 (50.6-72.7) | 54.6 (51.0-67.4) | 51.0 (47.7-67.1) | 61.2 (51.0-76.3) | 58.8 (50.5-74.7) | **0.037** | 0.193 | 0.087 | 0.297 |
| Hip (cm) | 70.9 (57.7-87.0) | 60.9 (55.4-85.3) | 53.9 (50.7-71.4) | 72.0 (56.4-89.8) | 69.4 (55.3-86.1) | 0.130 | 0.151 | 0.058 | 0.194 |
| Waist hip ratio | 0.86 (0.82-0.90) | 0.88 (0.85-0.92) | 0.92 (0.90-0.98) | 0.89 (0.83-0.94) | 0.89 (0.84-0.94) | 0.249 | 0.344 | 0.773 | 0.650 |
| Sum of skinfolds (mm) | 40.5 (25.6-74.8) | 32.7 (25.6-58.2) | 28.7 (24.5-83.4) | 44.8 (27.6-88.1) | 43.4 (27.0-84.6) | 0.086 | 0.152 | 0.054 | 0.168 |
| **Paternal measurements at follow-up** | | | | | | | | | |
| **Measurements** | **No diabetes**  **(n=162)** | **Type 1 diabetes (n=19)** | **Type 2 diabetes**  **(n=14)** | **Gestational**  **Diabetes**  **(n=115)** | **All diabetes**  **(n=148)** | **p1** | **p2** | **p3** | **p4** |
| Age (years) | 39.7 (36.2-45.2) | 36.8 (33.6-44.7) | 39.9 (37.0-41.6) | 43.1 (38.5-46.6) | 41.8 (37.5-45.9) | **0.002** | **0.003** | **0.001** | **0.021** |
| Height (cm) | 167.7 (163.9-173.5) | 168.7 (163.8-172.4) | 166.8 (163.9-171.6) | 168.4 (164.8-174.0) | 168.2 (164.6-173.2) | 0.594 | 0.381 | 0.496 | 0.716 |
| Weight (kg) | 72.0 (64.5-80.7) | 75.0 (60.8-84.9) | 65.9 (60.3-82.6) | 73.5 (67.1-84.8) | 73.2 (65.9-84.7) | 0.120 | **0.055** | **0.043** | 0.111 |
| BMI (Kg/m^2^) | 25.3 (23.2-27.6) | 25.9 (24.6-27.9) | 23.4 (22.2-29.1) | 25.8 (23.6-28.7) | 25.8 (23.5-28.7) | 0.223 | 0.120 | 0.063 | 0.114 |
| Overweight+obesity, n (%) | 89 (54.6) | 11 (57.9) | 6 (42.9) | 71 (61.2) | 88 (59.1) | 0.785 | 0.398 | 0.272 | 0.427 |
| Waist (cm) | 95.6 (89.3-101.5) | 97.0 (89.9-103.4) | 90.1 (81.8-107.7) | 97.1 (91.6-104.4) | 96.9 (90.1-104.4) | 0.223 | 0.097 | 0.062 | 0.158 |
| Hip (cm) | 98.3 (95.2-104.0) | 99.4 (93.5-103.0) | 94.3 (88.0-105.6) | 100.9 (95.3-106.4) | 100.2 (94.6-105.5) | 0.197 | **0.051** | 0.104 | 0.316 |
| Waist hip ratio | 0.96 (0.93-0.99) | 0.97 (0.92-1.01) | 0.95 (0.91-1.03) | 0.97 (0.93-1.00) | 0.97 (0.93-1.00) | 0.114 | 0.719 | 0.645 | 0.797 |
| Central obesity, n (%) (Waist hip ratio >0.9) | 148 (91.4) | 16 (84.2) | 11 (78.6) | 103 (88.8) | 130 (87.2) | 0.312 | 0.120 | 0.476 | 0.240 |
| Glucose intolerance (prediabetes+diabetes),  n (%) | 91 (55.8) | 12 (63.2) | 7 (50.0) | 80 (69.0) | 116 (67.4) | 0.542 | 0.674 | **0.026** | **0.055** |

Classification of pregnancies as non-diabetic and child’s birthweight in this group were based on maternal recall, diabetic pregnancies data was available in clinic records

Values are median (25^th^-75^th^ centile) for continuous variables n (%) for categorical variables

Offspring birth weight SD score was calculated for the group by adjusting for gestational age at delivery and gender.

Parental overweight-obesity (BMI>=25 Kg/m^2^), glucose intolerance in father (ADA criteria, pre-diabetes+diabetes)

p1: type 1 diabetes Vs no diabetes; p2: type 2 diabetes Vs no diabetes, p3: GDM Vs no diabetes, p4: All Vs no diabetes

**Table S4: Characteristics of Offspring of Diabetic mothers (ODM) and Offspring of Non- Diabetic mothers (ONDMs)**

|  | **All offspring (n=377)** | | | | | | | |
| --- | --- | --- | --- | --- | --- | --- | --- | --- |
|  | **Boys** | | | | **Girls** | | | |
|  | **n** | **ODM**  **(n=121)** | **n** | **ONDM**  **(n=103)** | **n** | **ODM**  **(n=79)** | **n** | **ONDM**  **(n=74)** |
| Age (years) | 121 | 8.0 (4.6-12.3) | 103 | 9.1 (5.6-13.0) | 79 | 8.4 (4.1-12.0) | 74 | 9.7 (6.8-12.9) |
| BMI (Kg/m^2^) | 118 | 16.0 (14.2-20.9) | 102 | 15.7 (14.1-18.5) | 79 | 16.8 (14.4-20.1) | 74 | 16.6 (14.0-20.1) |
| Overweight+obesity n (%) | 118 | 28 (23.7%) ** | 102 | 11 (10.8%) | 79 | 19 (24.1%) | 74 | 14 (18.9%) |
| Waist circumference (cm) | 119 | 57.8 (49.6-78.3) | 103 | 59.5 (51.0-72.4) | 79 | 61.2 (51.0-73.4) | 74 | 60.0 (50.3-74.8) |
| Central obesity n (%) | 118 | 52 (44.1) ** | 102 | 27 (34.2) | 79 | 29 (36.7) | 73 | 27 (37.0) |
|  | **< 10-year-old offspring (n= 212)** | | | | | | | |
|  | **n** | **ODM**  **(n=76)** | **n** | **ONDM**  **(n=55)** | **n** | **ODM**  **(n=43)** | **n** | **ONDM**  **(n=38)** |
| Age (years) | 76 | 5.5 (3.8-7.4) | 55 | 5.9 (4.7-8.2) | 43 | 4.6 (3.2-6.9) | 38 | 7.0 (3.9-8.7) |
| Overweight+obesity n (%) | 73 | 10 (13.7%) * | 54 | 1 (1.9%) | 43 | 9 (20.9%) | 38 | 4 (10.5%) |
| Central obesity n (%) | 73 | 27 (37.0) * | 54 | 11 (20.4) | 43 | 17 (39.5) | 37 | 11 (29.7) |
| Capillary fasting glucose (mmol/L) | 43 | 5.7 (5.4-5.9) *** | 35 | 4.9 (4.4-5.4) | 21 | 5.7 (5.4-5.9) *** | 25 | 4.9 (4.3-5.4) |
| Capillary fasting glucose > median n (%) | 43 | 30 (69.8%) *** | 35 | 8 (22.9%) | 21 | 17 (81%) *** | 25 | 7 (28%) |
| Capillary non-fasting glucose (mmol/L) | 30 | 5.6 (5.2-6.0) | 20 | 5.4 (4.6-6.1) | 21 | 5.5 (5.2-5.9) | 13 | 5.2 (4.9-6.2) |
| Capillary non-fasting glucose > median n (%) | 30 | 16 (53.3%) | 20 | 9 (47.4%) | 21 | 10 (47.6%) | 13 | 6 (46.2%) |
|  | **≥10-year-old offspring (n=165)** | | | | | | | |
| **Parameter** | **n** | **ODM**  **(n=45)** | **n** | **ONDM**  **(n=48)** | **n** | **ODM**  **(n=36)** | **n** | **ONDM**  **(n=36)** |
| Age (years) | 45 | 13.8 (11.9-18.6) | 48 | 13.6 (11.7-18.2) | 36 | 12.1 (10.8-17.9) | 36 | 12.9 (11.3-17.4) |
| Central obesity n (%) | 45 | 25 (55.6) * | 48 | 16 (33.3) | 36 | 12 (33.3) | 36 | 16 (44.4) |
| Fasting glucose (mmol/L) | 45 | 5.4 (5.2-5.6) ** | 48 | 5.0 (4.8-5.3) | 36 | 5.3 (4.9-5.5) ** | 36 | 4.9 (4.7-5.2) |
| 30 min glucose (mmol/L) | 42 | 8.6 (7.7-9.3) ** | 47 | 7.9 (7.2-8.7) | 35 | 8.5 (7.2-9.2) * | 35 | 7.4 (6.9-8.2) |
| 120 min glucose (mmol/L) | 41 | 6.4 (5.6-8.1) * | 47 | 6.1 (5.5-7.4) | 35 | 6.5 (5.6-7.4) * | 35 | 5.9 (4.9-6.6) |
| HbA1C (mmol/mol) | 41 | 36 (33-39) * | 45 | 34 (32-37) | 31 | 37 (33-39) | 35 | 34 (33-38) |
| HbA1c (%) | 41 | 5.4 (5.2-5.7) * | 45 | 5.3 (5.1-5.5) | 31 | 5.5 (5.2-5.7) | 35 | 5.3 (5.2-5.6) |
| Prediabetes n (%) | 43 | 19 (42.2) * | 48 | 11 (22.9) | 35 | 11 (30.6) | 36 | 6 (16.7) |
| Diabetes n (%) | 3 | 3 (6.6) | 0 | 0 (0) | 1 | 1 (2.7) | 0 | 0 (0) |

ODM: Offspring of diabetic mothers, ONDM: Offspring of non-diabetic mothers, HOMA: Homeostatic model for assessment, HDL: high density lipoprotein

Central obesity: waist to height ratio>0.5, Prediabetes: IFG+IGT+Both, Diabetes: Known + newly diagnosed, Disposition Index: [Log (Insulinogenic index) + Log (Matsuda index)].

*:p<0.05, **:p<0.01, ***:p<0.000

**Table S5: Metabolic-endocrine characteristics of Hyperglycemic vs. Normoglycemic ODMs (>=10 years) based on OGTT**

| **Measurements** | **Hyperglycemic**  **(n=31)** | **Normoglycemic**  **(n=47)** | **p1** | **p2** |
| --- | --- | --- | --- | --- |
| Boys n (%) | 20 (64.5) | 23 (48.9) | **--** | **--** |
| Age (years) | 14.2 (12.0-17.9) | 12.2 (11.1-16.8) | 0.526 | -- |
| BMI (Kg/m^2^) | 21.8 (18.8-26.6) | 19.4 (16.8-22.9) | **0.023** | -- |
| Fasting glucose (mmol/L) | 5.61 (5.40-5.76) | 5.22 (5.05-5.33) | **0.000** | **0.000** |
| 30 min glucose (mmol/L) | 9.05 (8.33-9.72) | 8.11 (7.22-8.77) | **0.000** | **0.000** |
| 120 min glucose (mmol/L) | 8.00 (6.72-9.22) | 6.22 (5.47-6.63) | **0.000** | **0.000** |
| HbA1c (%) | 5.6 (5.3-5.8) | 5.3 (5.1-5.5) | **0.004** | **0.020** |

Values in median (25^th^-75^th^ percentile), p1: adjusted for age and gender except for age, p2: adjusted for age, gender and child’s BMI except for age

ODM: Offspring of diabetic mothers, ONDM: Offspring of non-diabetic mothers, OGTT: Oral glucose tolerance test

**Figure S1: Agreement between venous plasma glucose and capillary glucose in parents**

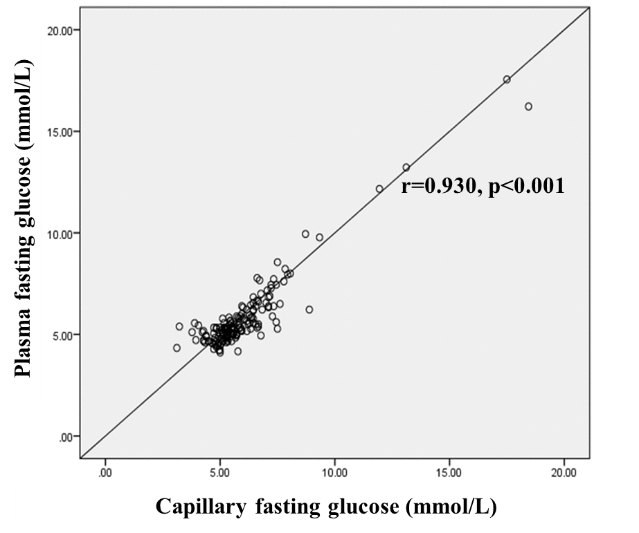

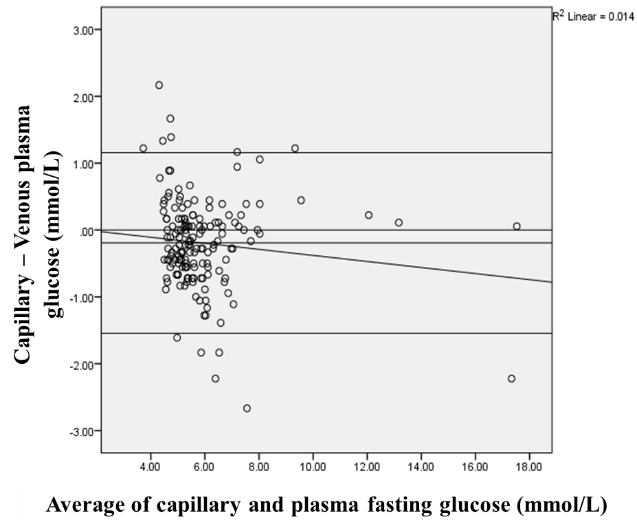

**Figure 1b**

**Figure 1a**

1 a: Correlation between capillary fasting glucose and plasma fasting glucose in parents

1 b: Bland Altman Plot shows distribution of differences between capillary plasma glucose measured on glucometer and venous blood glucose vs. mean of the two measurements. A negative bias of 0.19 mmol/L is represented by line A.

Figure S2: Flow diagram of the study

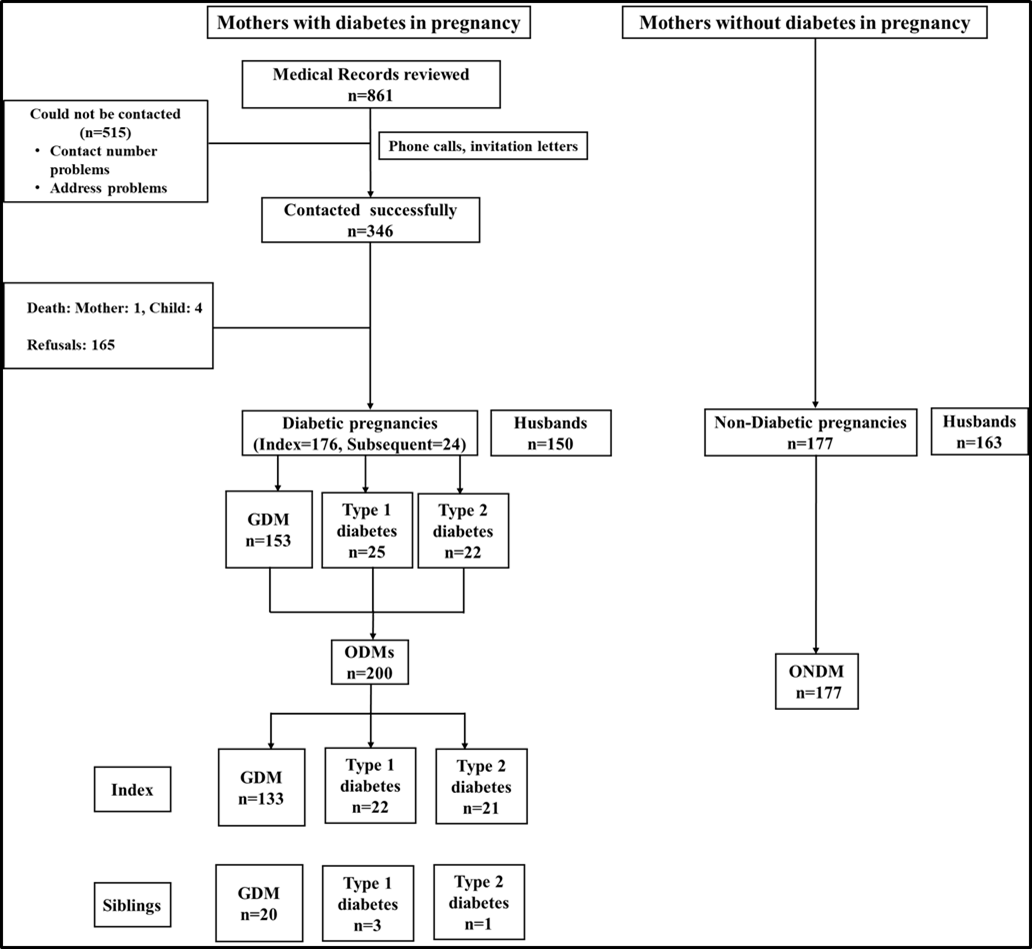

Figure shows flow diagram of the study. Women attending pregnancy diabetes clinic were contacted to participate in the follow up of their children. Families who consented were studied. Control children whose mothers were not diabetic during pregnancy were studied along with their parents.

ODM: Offspring of diabetic mothers, ONDM: Offspring of non-diabetic mothers, GDM: Gestational Diabetes Mellitus

**Figure S3- Body size and glycemic measurements in ODM and ONDM**

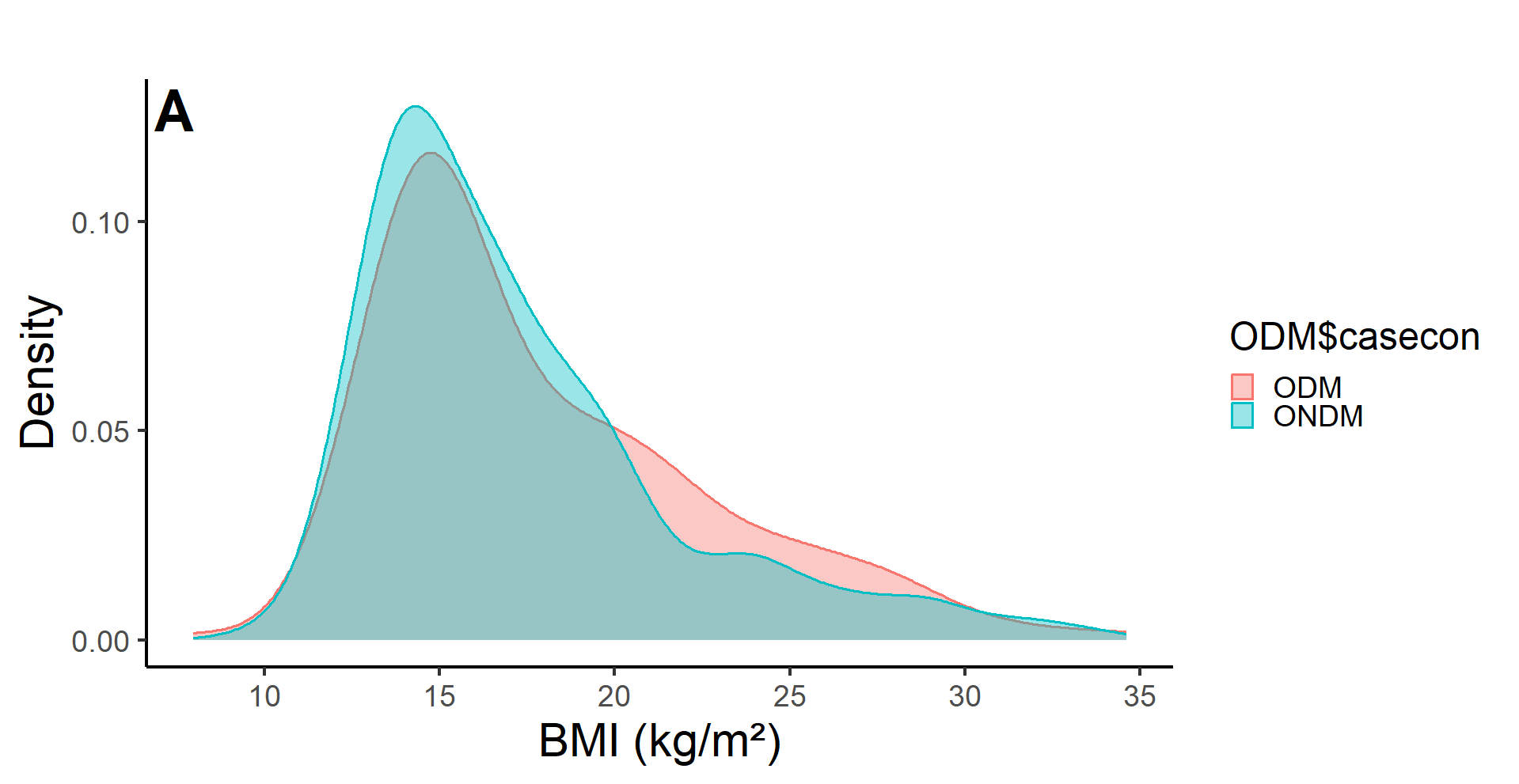

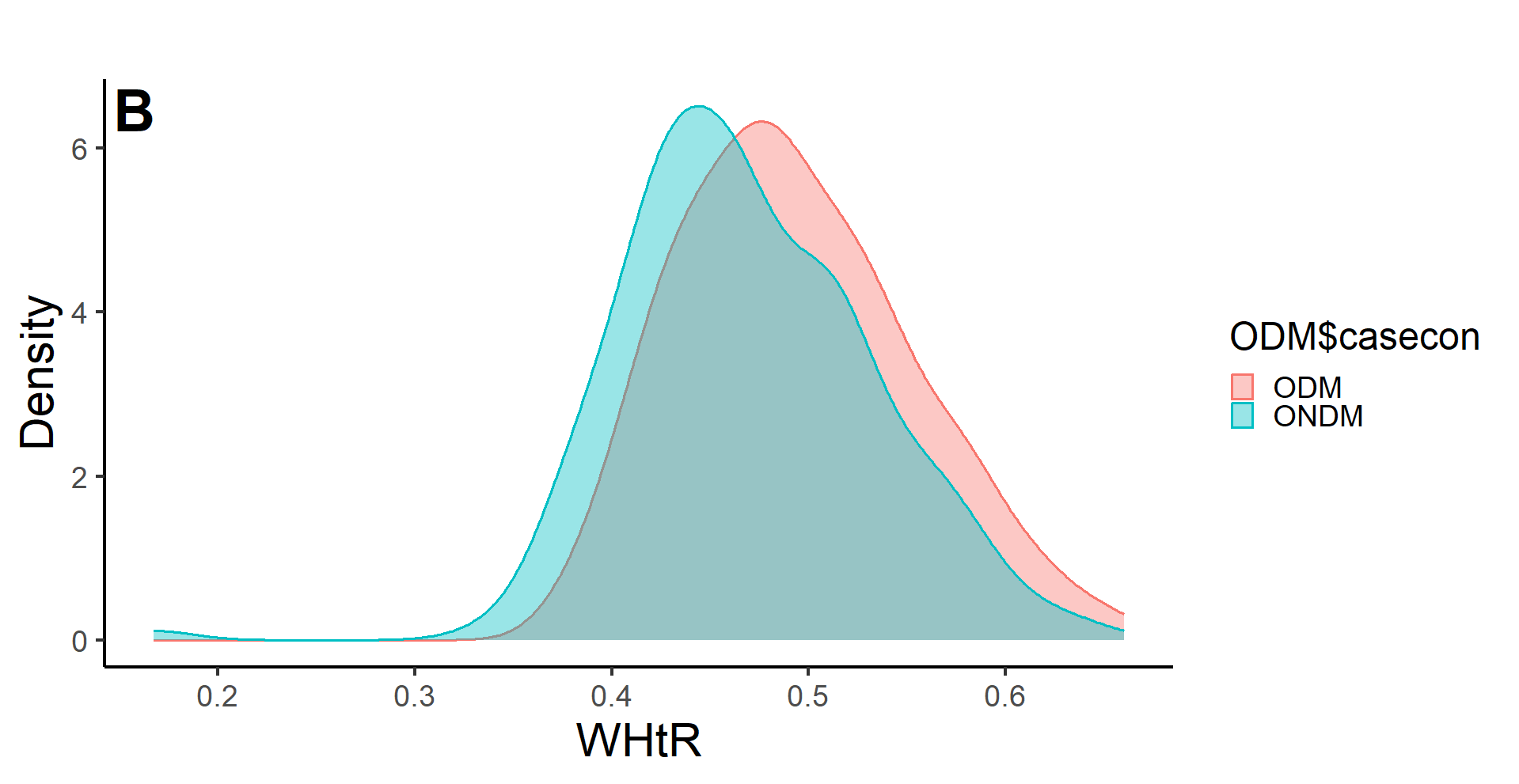

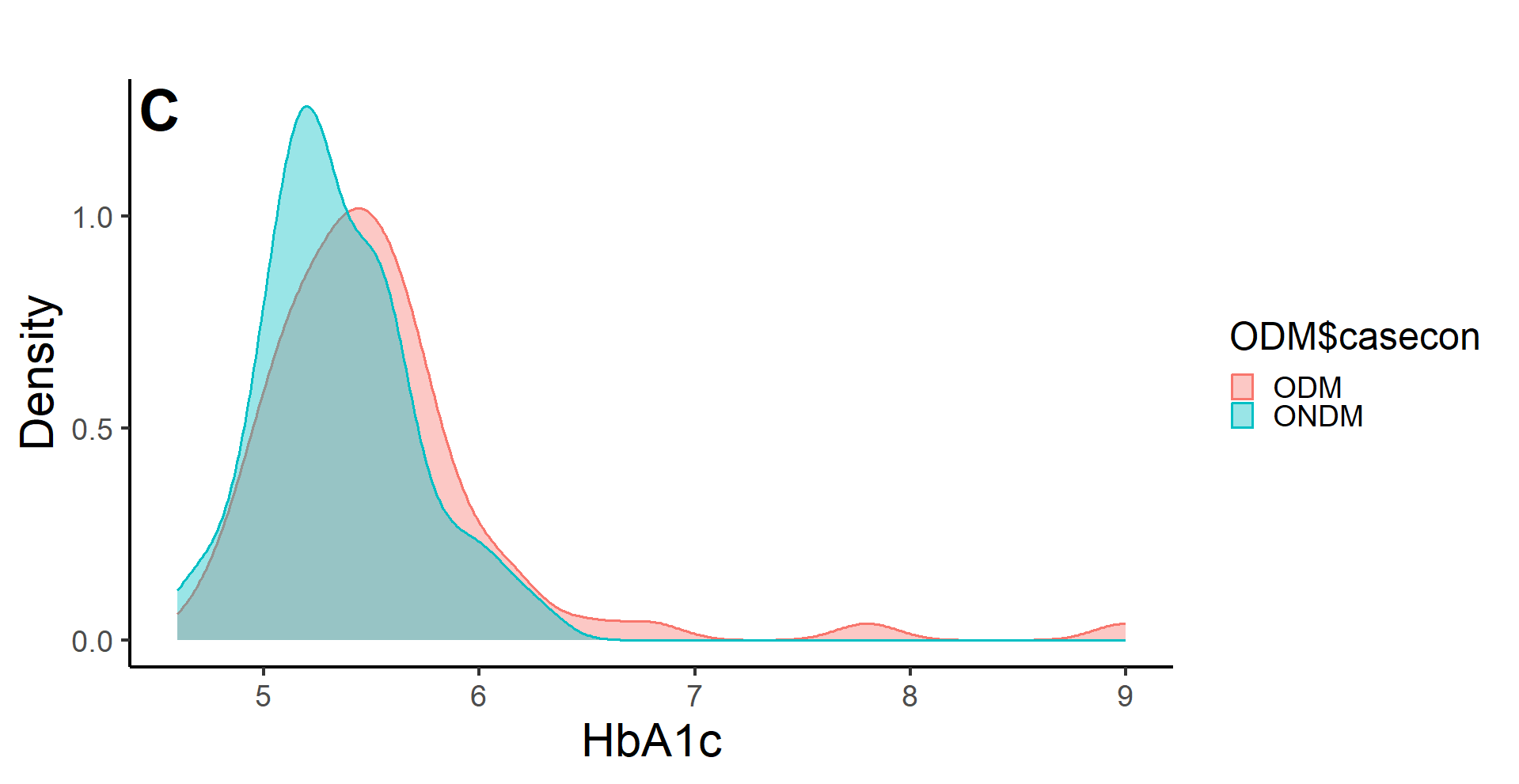

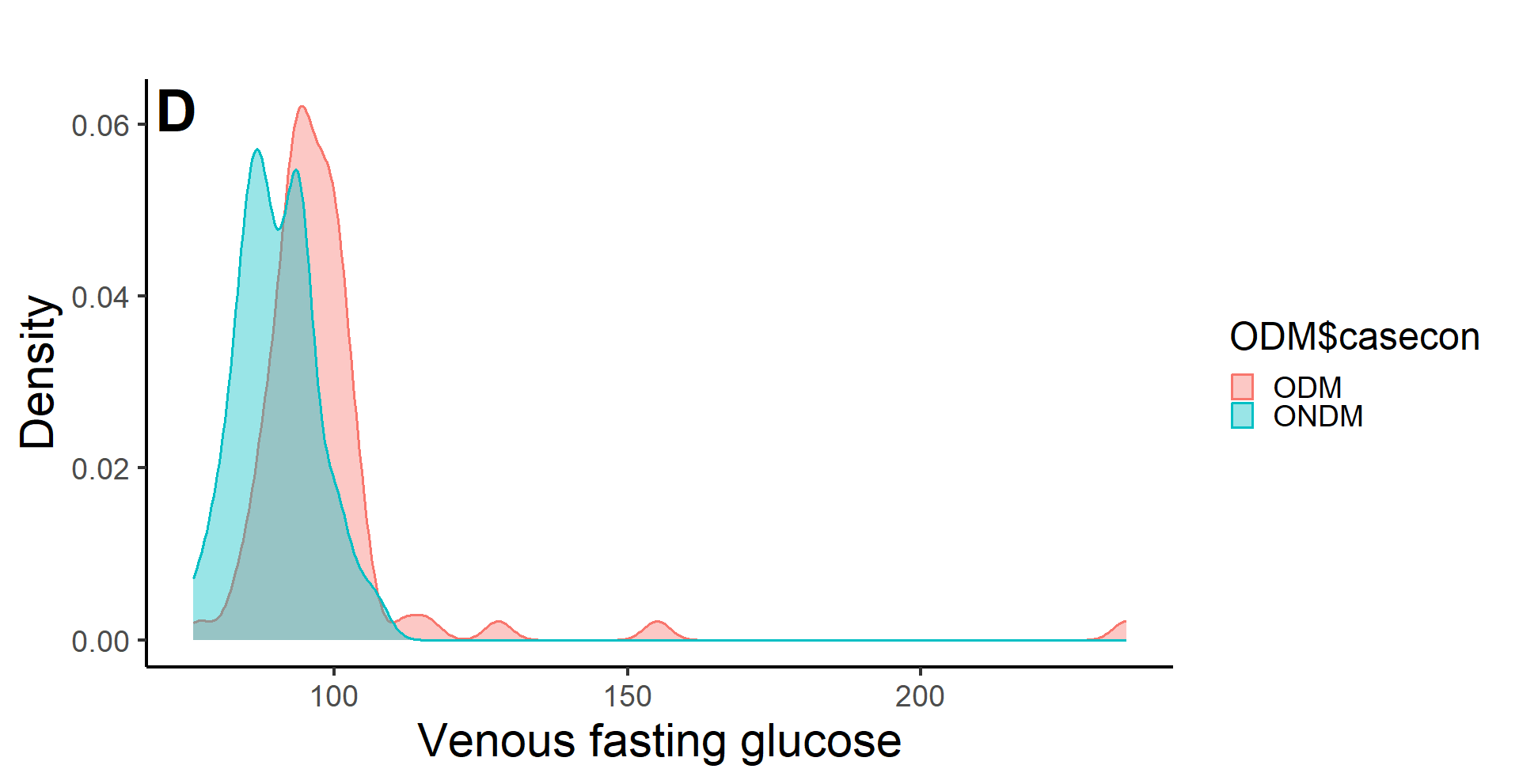

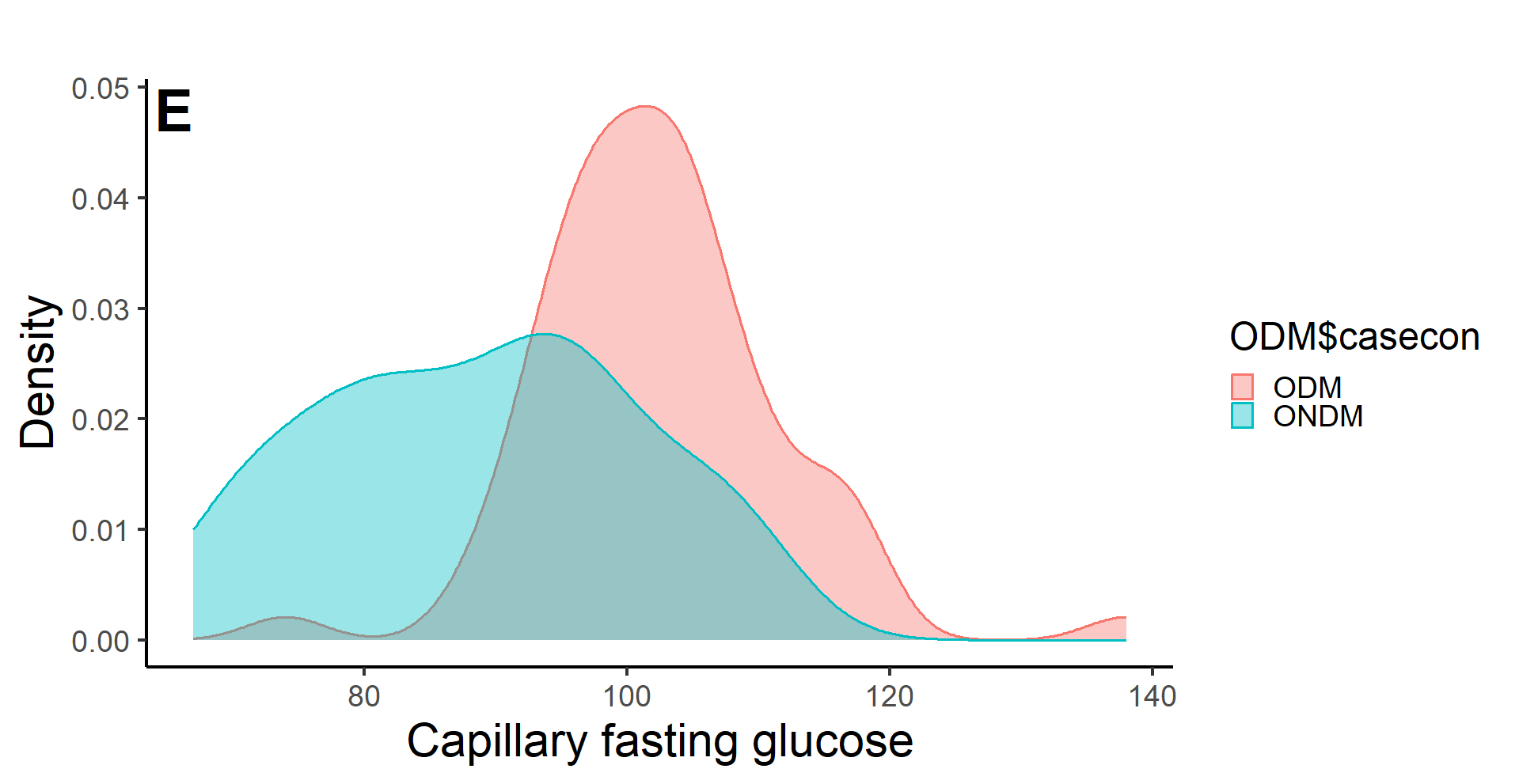

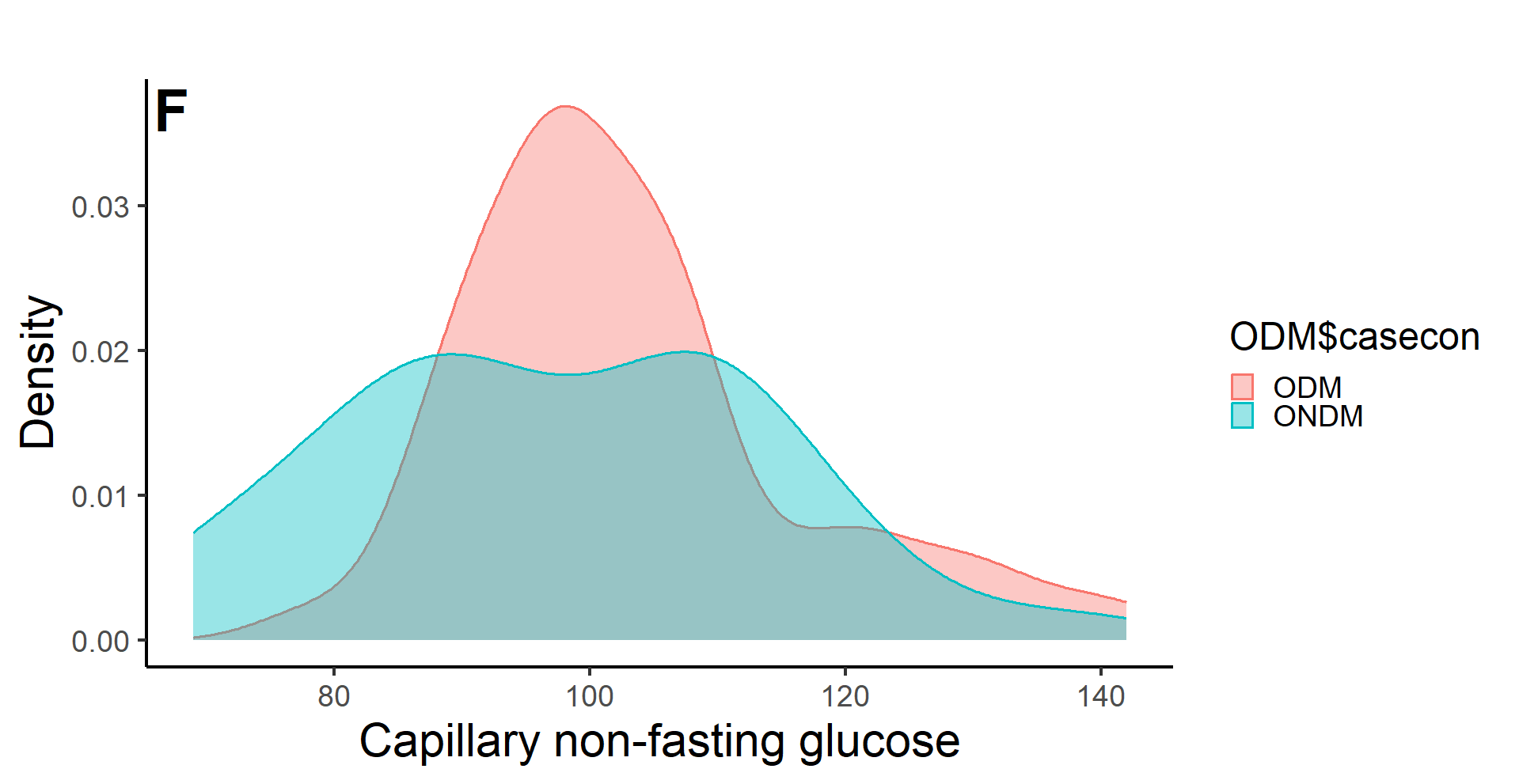

Figure shows distribution of body size and glycemia in the ODM-offspring of mothers with diabetes in pregnancy, indicated in pink colour and ONDM-offspring of mothers without diabetes in pregnancy, indicated in blue colour.

**Figure S4: Overweight + obesity in offspring according to parental size**

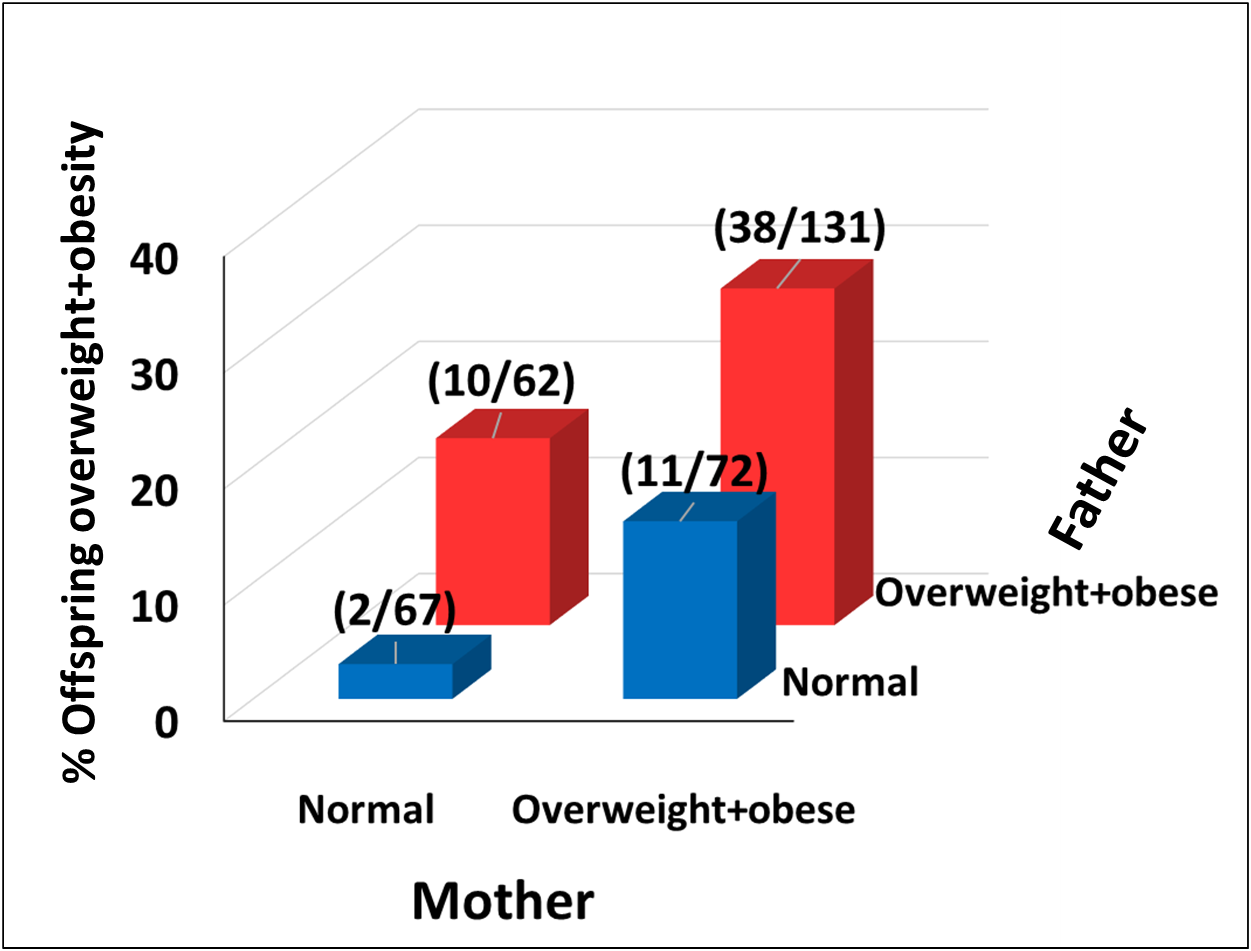

Figure shows association between parental and offspring overweight-obesity, data from diabetic and non-diabetic pregnancies is combined. Offspring: overweight-obesity was classified using WHO criteria (SD scores by MGRS 2007 for <=18y and WHO criteria BMI>=25 Kg/m^2^ for >18 years). Overweight-obesity in parents ((WHO criteria, BMI >=25 Kg/m^2^) was measured at follow up.

ODM: Offspring of mothers with diabetes, ONDM: Offspring of mothers without diabetes, IOTF: International Obesity Task Force, WHO: World Health Organization

**Figure S5: Obesity and glycemia in the parents and offspring at follow up according to maternal type of diabetes in pregnancy**

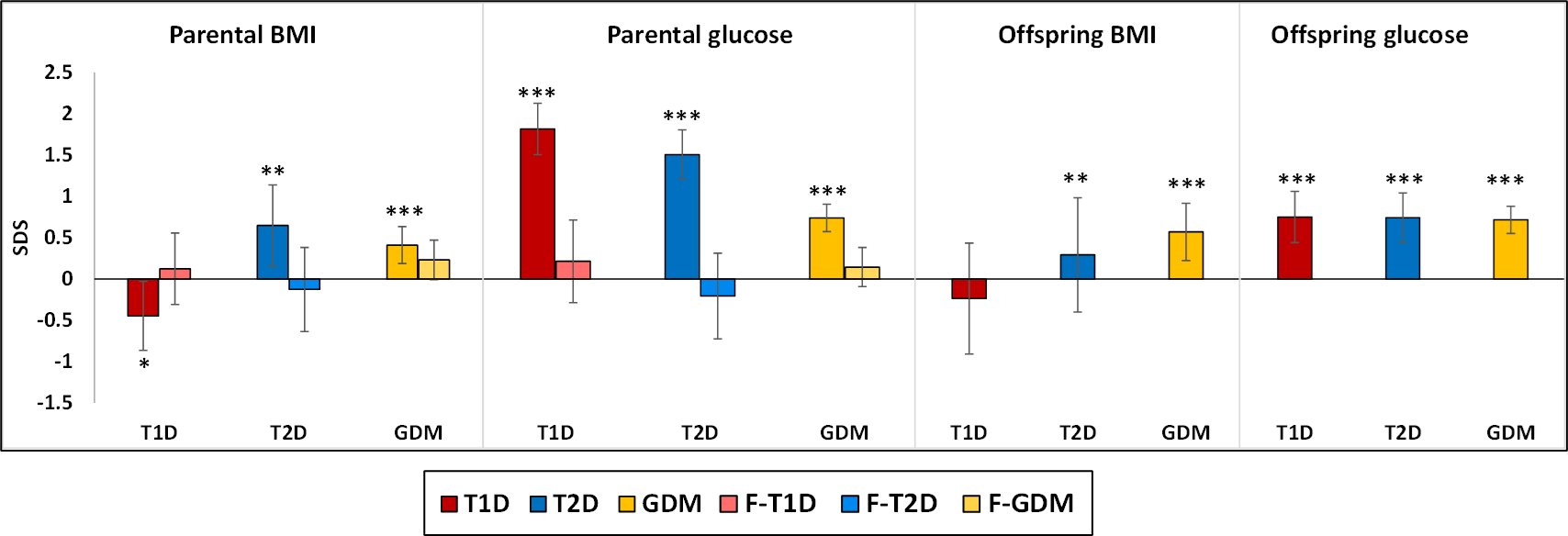

Figure shows parental and offspring characteristics by type of maternal diabetes. T1D: Type 1 diabetes, T2D: Type 2 diabetes, GDM: Gestational diabetes mellites, F-T1D: father of offspring born to mother with type 1 diabetes, F-T2D father of offspring born to mother with type 2 diabetes, F-GDM: father of offspring born to mother with gestational diabetes mellitus. Bar height represents mean difference in the respective measurements; zero line depicts measurements for the non-diabetic category.
